# PREPRINT: Cluster analysis of ME/CFS symptoms in DecodeME reveals two subgroups and a link to onset type

**DOI:** 10.64898/2026.06.29.26356818

**Authors:** Christa St-Jean, Joshua J. Dibble, Chris Ponting, Regina Prigge

## Abstract

**Background:** Myalgic encephalomyelitis/chronic fatigue syndrome (ME/CFS) is a debilitating, often infection-triggered illness with no cure and no effective treatment. Marked symptom heterogeneity hampers diagnosis, disease management, and trial design. Using phenotype data from the world’s largest ME/CFS cohort, this study aimed to identify groups of patients with similar symptom profiles using cluster analysis, to assess the association between cluster membership and onset type, and to explore genetic associations with cluster membership.

**Methods:** This study included 19,019 DecodeME participants, ages 16 and over, with ME/CFS in the UK, from 2022-2024. We performed a *k*-modes cluster analysis of individuals based on similar symptoms. Cluster metrics identified the optimal number of clusters, which were characterised and compared. A sex-stratified subgroup analysis explored differences between clusters among males and females. The association between ME/CFS onset type (infectious, non-infectious, or unknown) and cluster membership was assessed with logistic regression models, adjusting for sex, age, deprivation, and ethnicity. Genetic associations with cluster membership were assessed using a genome-wide association study.

**Results:** We identified two clusters in our study population: a high symptom burden cluster (HSBC; 57% of participants) and a lower symptom burden cluster (LSBC; 43%). The HSBC was characterised by higher prevalence of symptoms across all domains, more comorbidities, and greater illness severity. Individuals with infectious and unknown onset had 1.24 times (95% CI: 1.15-1.35) and 1.30 times (95% CI: 1.18-1.43) higher adjusted odds of HSBC membership relative to non-infectious onset, respectively. A similar pattern was observed in the sex-stratified analyses, although it showed an overall higher symptom prevalence for females and a higher proportion of females in the HSBC compared to males. No genetic variant was significantly associated with cluster membership.

**Conclusions:** This large-scale cluster analysis of DecodeME symptom data reinforces that ME/CFS is a heterogeneous condition with clinical subtypes. The identification of symptom-based phenotypes, along with sex-based differences in symptom burden and cluster characteristics, highlights the importance of incorporating symptom burden and sex in future research, clinical decision-making, and public health strategies. Tailoring future interventions to these subgroups could enhance patient management and improve outcomes.

**Lay Summary:** Myalgic encephalomyelitis/chronic fatigue syndrome (ME/CFS) is a serious illness affecting tens of millions of people worldwide. Severe fatigue, cognitive symptoms (problems with thinking, concentration, or memory), and post-exertional malaise (a prolonged worsening of symptoms, or new symptoms, after even minor mental or physical effort) are core ME/CFS symptoms, though individuals can experience a wide variety of other symptoms. Some of these symptoms overlap with other illnesses, making diagnosis difficult. Determining if there are subgroups of individuals with ME/CFS that share similar symptoms can help doctors and researchers better understand the different ways that ME/CFS affects people, which could lead to improved treatments, and can help direct research into its underlying causes. The present study analysed ME/CFS symptom data from a large UK-based sample using a computer program that sorts individuals into groups purely based on their shared symptoms. Two groups were consistently found: one where people had a wide range of ME/CFS symptoms, and another where people had fewer symptoms. Females tended to have more symptoms overall and so were more likely than males to be in the group with widespread symptoms. Many individuals report that an infection triggered their ME/CFS symptoms, but there are other possible triggers (e.g., physical or emotional trauma). The study found a link between ME/CFS being triggered by an infection and having more symptoms. These findings indicate that ME/CFS is not a “one-size-fits-all” disease, that we need more research to understand what causes ME/CFS, and that healthcare should be tailored to each person’s needs.

## Introduction

Myalgic encephalomyelitis/chronic fatigue syndrome (ME/CFS) is a chronic, debilitating neurological condition characterised by severe fatigue that is not alleviated by rest (typically at least 6 months in duration), cognitive impairment, and post-exertional malaise (PEM; a prolonged worsening of symptoms, or new symptoms, after mental or physical exertion) (1). Although ME/CFS primarily affects the nervous and immune systems, individuals with ME/CFS can experience a multitude of symptoms affecting many other body systems (e.g., gastrointestinal, genitourinary, respiratory), often fluctuating in type and severity throughout the course of illness and causing a significant reduction in functional capacity and quality of life (2). ME/CFS symptoms place a substantial burden on patients, their loved ones, and their carers.

The exact causes of ME/CFS are uncertain. A recent review found that 50%-80% of individuals with ME/CFS began symptoms at the time of or following an infectious disease, suggesting possible immune system dysfunction at disease onset (1). The review also found evidence from multiple studies of a potential genetic link. To investigate a potential genetic component, the world’s largest cohort of individuals with ME/CFS was formed as part of the DecodeME study, which was co-produced by researchers, ME/CFS patients, their carers, ME/CFS charities, industry professionals, and members of the public (3,4).

While preliminary results of the DecodeME genetic study support a heritable component to ME/CFS (4), many knowledge gaps remain regarding the causes of ME/CFS, and how to diagnose and manage the disease. A recent James Lind Alliance Priority Setting Partnership (JLA PSP) was formed to identify the Top 10 research priorities for ME/CFS stakeholders to direct future research and funding (5). The present study aims to address research priority #5: “Are there different types of ME/CFS linked to different causes and how severe it becomes? Do different types of ME/CFS need different treatments or have different chances of recovery?” (6). The JLA PSP emphasises the wide variety of symptoms and variation in terms of which symptoms cause individuals the most disability, suggesting that future research and disease management would benefit greatly from ME/CFS being categorised into subgroups of individuals with similar symptoms, prognosis, or underlying biology (6).

The aim of this study is to identify groups of patients with similar symptom profiles using cluster analysis, to determine whether cluster characteristics differ by sex, to assess the association between cluster membership and onset type, and to explore genetic associations with cluster membership.

## Methods

### Study Population

In this cross-sectional study, we analysed phenotype data from DecodeME. Over 26,000 UK residents aged 16 years and older and with a self-reported clinical diagnosis of ME/CFS were recruited between September 2022 and November 2023. Participants were enrolled through a combination of charity-led mail-outs, digital marketing initiatives, traditional and social media campaigns, and person-to-person recruitment (7). We obtained de-identified data from the 23,029 participants who agreed to share their data with other researchers. Exclusion criteria included: a self-diagnosis of ME/CFS, not meeting either the Canadian Consensus Criteria (CCC) or Institute of Medicine/National Academy of Medicine (IOM/NAM) case definitions for ME/CFS, and hospitalisation from COVID-19 and/or heart or lung damage due to severe COVID-19 in those individuals who reported they had a COVID-19 infection at the onset of their ME/CFS symptoms (7). Additionally, individuals whose sex at birth was indicated as ‘intersex’ or ‘prefer not to say’ were excluded from the cluster analysis due to very low numbers of individuals in those categories, prohibiting subgroup or sensitivity analyses. Individuals without an active study status or who had withdrawn from the study were also excluded, as was one individual whose very high reported age (123 years) was determined to be an extreme outlier. The very small number of participants with missing information on ethnicity (n = 51, 0.26%) and comorbidities (≤ 0.21% missing data for each of the 34 comorbidities) were excluded from the analysis.

This present study was covered under the existing ethical approval of the DecodeME study which received a favourable opinion from the North West – Liverpool Central Research Ethics Committee (21/NW/0169).

All data cleaning, preprocessing, exploration, and analyses were performed using R version 4.1.2 (8).

### ME/CFS Symptoms

This analysis is based on 67 binary symptom variables, ascertained through the DecodeME study baseline questionnaire (9) which was developed from the CCC and IOM/NAM diagnostic criteria by individuals with lived experience of ME/CFS (3). Variables assessing the presence of fatigue and PEM were excluded from the cluster analysis as they are present in 100% of participants (required to meet both CCC and IOM/NAM case definitions). Similarly, the following four additional symptom variables that are present in over 90% of individuals were excluded: 1) unrefreshing sleep, 2) confusion or ‘brainfog’, 3) finding it hard to concentrate, and 4) a variable assessing the type of fatigue experienced (physical, mental, both, or none). These symptoms were excluded because variables with minimal variability are not useful for discriminating between clusters and may reduce clustering performance and introduce bias (10).

### Covariates

Participants’ outward postcodes were used as a proxy for socioeconomic status by linking them to Index of Multiple Deprivation (IMD) deciles via the 2019 Harmonised IMD dataset for UK countries (11) and the February 2025 Office for National Statistics postcode directory dataset for the UK (ONSPD) (12). Each postcode was assigned to its most common lower-layer super output area and matched to an IMD decide, which was then converted to a quintile to simplify analysis and enhance statistical power. For two individuals (0.01%), their postcodes did not map to an LSOA. These values were imputed using mode imputation. Due to small numbers of individuals in ethnic minority groups, the levels ‘Asian or Asian British’, ‘Black, African, Caribbean, or Black British’, ‘Mixed or multiple ethic groups’, and ‘Other ethnic group’ were collapsed into one ‘Ethnic minority’ level. Additionally, due to small numbers of individuals having COVID-19 at ME/CFS onset, the three infection categories (glandular fever, COVID-19, and other infection) were collapsed into an ‘Infectious onset’ category. Individuals who answered “Don’t know” to whether they had an infection at the time of, or just prior, to ME/CFS symptom onset are defined as “unknown” onset type. In this study, males and females are defined by sex at birth.

### Statistical Analyses

#### Cluster Analysis

We conducted *k*-modes clustering to identify subgroups of participants with similar symptom profiles using the klaR() package in R (13). The *k*-modes algorithm was chosen for its ability to handle categorical data. Additionally, a recent study examining different clustering methods used with a dataset of 69 binary variables found that the *k*-modes algorithm consistently performed better than latent class analysis, agglomerative hierarchical clustering, and *k-*medoids clustering, based on a variety of clustering metrics (14). To determine the optimal number of clusters (*k*) we generated elbow plots based on the within-cluster dissimilarity across a range of *k* values (*k=2* to *k=10)*, identifying the point at which additional clusters provided minimal improvement in model fit. This selection was further validated using silhouette analysis, including both graphical inspection of silhouette plots and evaluation of average silhouette widths, to assess the cohesion and separation of clusters. The procedure was repeated for the total sample and for the sex-stratified subsamples to capture potential differences in cluster characteristics between females and males. As the number of males was substantially smaller than the number of females in the sample, we performed a bootstrap analysis (100 replicates) to assess the stability of the sex-specific clusters. Cluster stability was measured using the Adjusted Rand Index (ARI) (15).

#### Cluster Characterisation

Differences in baseline characteristics between clusters were assessed using a two-sample t-test for the age variable, and Chi-squared or Fisher’s exact tests for the categorical variables, with Bonferroni correction (16) applied for multiple testing.

We evaluated the distribution of each condition across clusters using two complementary measures. First, we calculated the Adjusted Relative Frequency (ARF) (14) to quantify the extent to which a condition was over- or under-represented in a cluster compared with its prevalence in the total cohort. An ARF value of 1 indicates equal prevalence, while values above or below 1 reflect over- and under-representation, respectively. Second, we assessed statistical significance of these differences using Fisher’s exact test (two-sided, α = 0.05), applying a Bonferroni correction to adjust for multiple testing. Statistically significant ARF values were subsequently visualized using a bubble heat map to facilitate comparison across clusters.

To determine which symptoms were drivers of cluster separation, we calculated effect sizes for each symptom using Cohen’s *h* for both the total sample and sex-stratified clusters. Cohen’s *h* values of 0.5 were used to indicate moderate effect, and 0.8 to indicate large effect (17). To formally test whether the between-cluster difference for each symptom varied by sex, we fitted symptom-specific logistic regression models including a cluster-by-sex interaction term in the total sample.

#### Inferential Analysis

To assess whether symptom–cluster associations differed by sex, we conducted symptom-specific binomial logistic regression analyses in the total sample, with each binary symptom as the outcome and cluster, sex, and a cluster-by-sex interaction term as predictors. The interaction term tested whether the difference in symptom prevalence between the high- and low-symptom burden clusters varied significantly between males and females; false discovery rate correction was applied across symptoms.

To examine the association between onset type and cluster membership, we conducted univariable and multivariable logistic regression analyses for the total sample and sex-stratified subgroups. The multivariable models adjusted for age, sex (in the total sample), ethnicity, and IMD decile. We evaluated the linearity of age with the log-odds of the outcome by comparing models using continuous, categorical (10-year bins), and spline-transformed age, selecting the best-fitting approach based on AIC, likelihood ratio tests, and plots of predicted probabilities.

Model selection and interaction effects were explored using the glmulti() function in R (18), which compared all possible models and pairwise interactions based on AIC. We assessed model diagnostics by examining residual deviance, Cook’s distance, leverage, and deleted residuals to detect influential points, and used variance inflation factors (VIF) to check for multicollinearity among independent variables.

#### Genome-Wide Association Study

Genetic data from DecodeME participants was extracted from the jointly imputed case-control dataset for GWAS-1 (4). For this analysis, we retained only participants who were included in DecodeME GWAS-1 and had matching questionnaire data, leading to a dataset with 15,328 participants. Of these, 2,401 participants were allocated to the LSBC, and 3,264 to the HSBC. We used REGENIE (19) for genome-wide association testing using the same parameters as for GWAS-1 (4) for autosomal variants with minor allele frequencies ≥ 1% and imputation INFO scores ≥ 0.9. For covariates, we included sex, principal components 1-20 calculated for GWAS-1, and genotyping batch. Effects due to population stratification were well controlled (λ = 1.044).

## Results

### Study Population

The study population included 19,019 participants aged 16 years and over (Figure 1). Most participants were female (85%) and of white ethnicity (97%). The average age of the sample was 48.7 years (sd = 14.5) (Supplemental Table 1).

**Figure 1.**
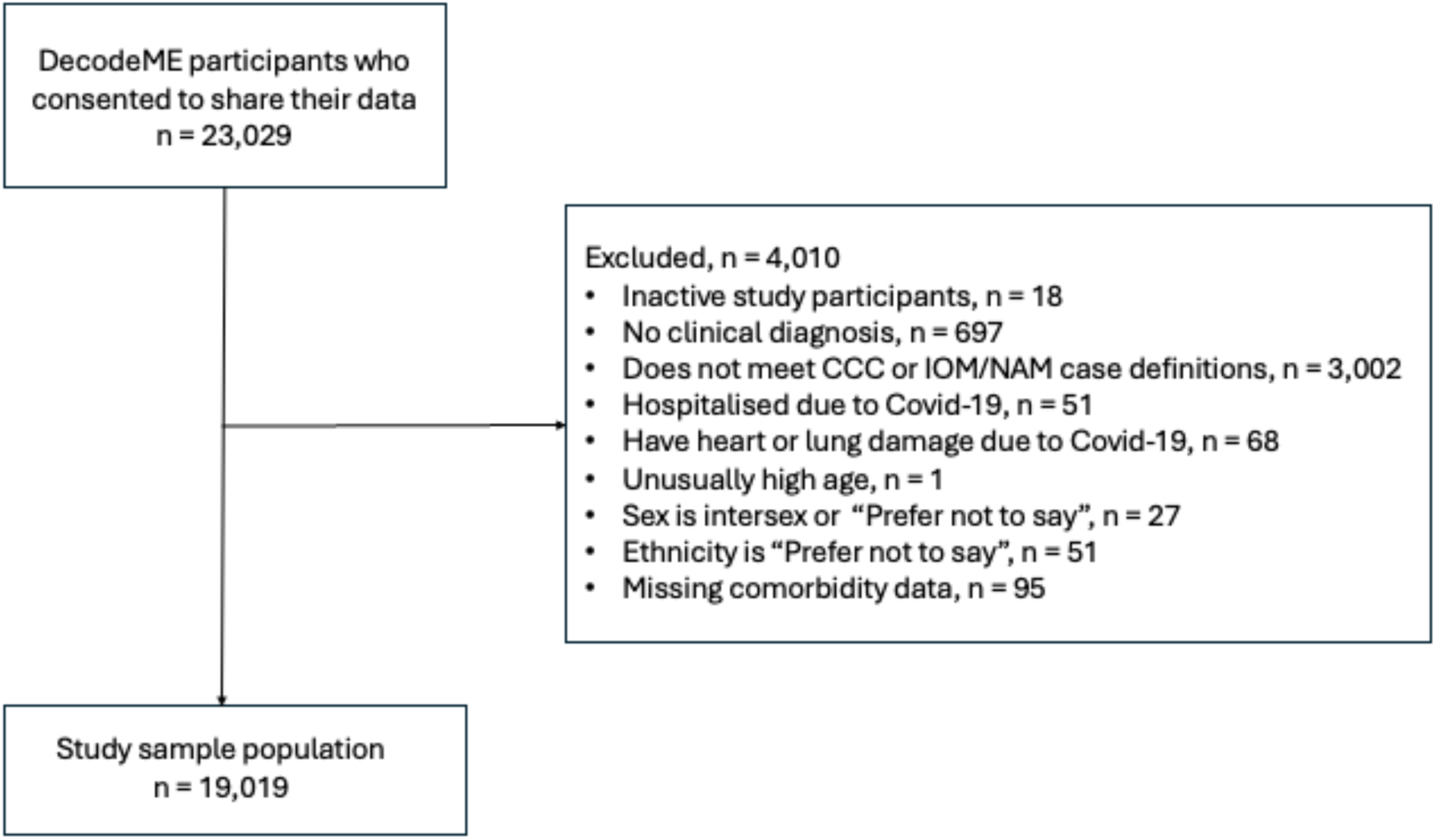
Flow diagram of sample selection.

### Cluster Analysis

Analysis of the elbow plot for the total sample showed a two-cluster solution was optimal (Supplemental Figures 1A, 1B, and 2).

Characterisation of the clusters indicated that the primary distinction between subtypes was overall symptom burden, separating participants into a high symptom burden cluster (HSBC) and a lower symptom burden cluster (LSBC). (Supplemental Table 2). The HSBC contained more individuals (n = 10,849, 57%) than the LSBC (n = 8,170, 43%), and a higher proportion of females (89% vs 80% in the LSBC, p < 0.001). The mean age of individuals in the HSBC was lower than that of the LSBC (46.7 years versus 51.5 years, p < 0.001). There was no difference in ethnicity between clusters. Compared to the LSBC, there were slightly higher proportions of individuals in the HSBC living in areas of higher deprivation (e.g., 22.6% vs 19.3% for Quartile 1, and 24.9% vs 23.7% for Quartile 2, p < 0.001). Approximately 61% of individuals in both clusters reported an infectious onset. The HSBC had slightly fewer individuals with non-infectious onset (15.5% vs 17.5%, p < 0.001) and more individuals with unknown onset (23.4% vs 21.2%, p < 0.001). Over half of participants in both clusters had ME/CFS for ≥10 years, and about 10% for <3 years. Severe and very severe illness were both around three times more common in the HSBC than in the LSBC (severe: 18% vs 6%, p < 0.001; very severe: 1.1% vs 0.3%, p < 0.001). The HSBC also had more individuals reporting worsening illness (22.7% vs 11.4%, p < 0.001) and fewer reporting improvement (1.2% vs 4.3%, p < 0.001). Comorbidities were generally more prevalent in the HSBC, while cancer was the only comorbidity more prevalent in the LSBC (p < 0.001).

Overall, individuals in the HSBC showed a broad overrepresentation of symptoms across nearly all domains, while those in the LSBC showed corresponding underrepresentation (Figure 2). The most prominent differences were observed for flu-like symptoms (sore throat, fever or chills, swollen glands, and viral infections) and sensory sensitivities (light, noise, and smell). Marked differences were also seen for musculoskeletal symptoms (muscle pain, stiffness, weakness, and pain spread), as well as cognitive and coordination-related symptoms, such as slow thinking, memory problems, and balance difficulties. In addition, autonomic and neuroendocrine symptoms (e.g., dizziness, palpitations, temperature regulation problems, excessive sweating) were more common in the HSBC, along with mood and energy-related symptoms, such as mood swings and racing thoughts. In this analysis, 58 out of the 67 symptoms, all of which are more prevalent in the HSBC, have moderate-to-large effects on cluster separation, based on Cohen’s *h* ≥ 0.5 (Table 1).

**Figure 2.**
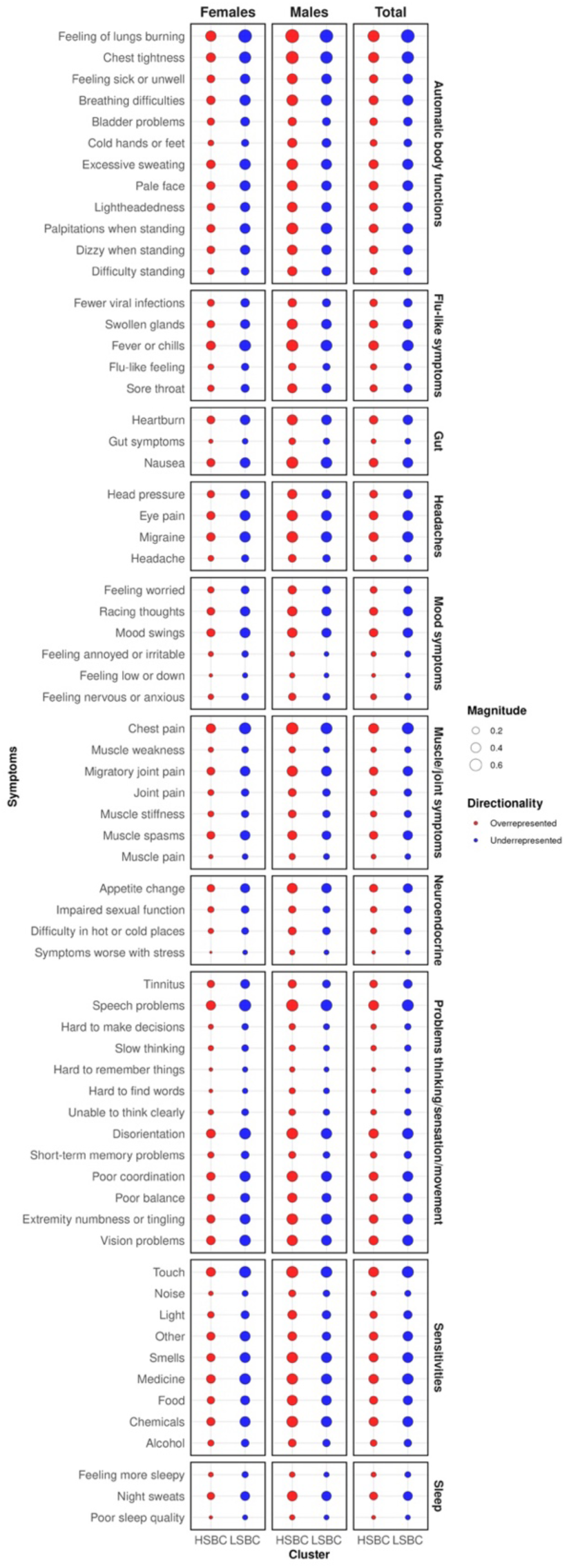
Bubble heat map showing symptom over- and under-representation in each cluster for both the total sample and for females and males. The magnitude represents the level of over- (red bubbles) or under- (blue bubbles) representation, where the larger the magnitude (larger the bubble), the greater the degree to which the symptom is over- or under- represented in the cluster, relative to the prevalence of the symptom in the entire sample.

**Table 1.**
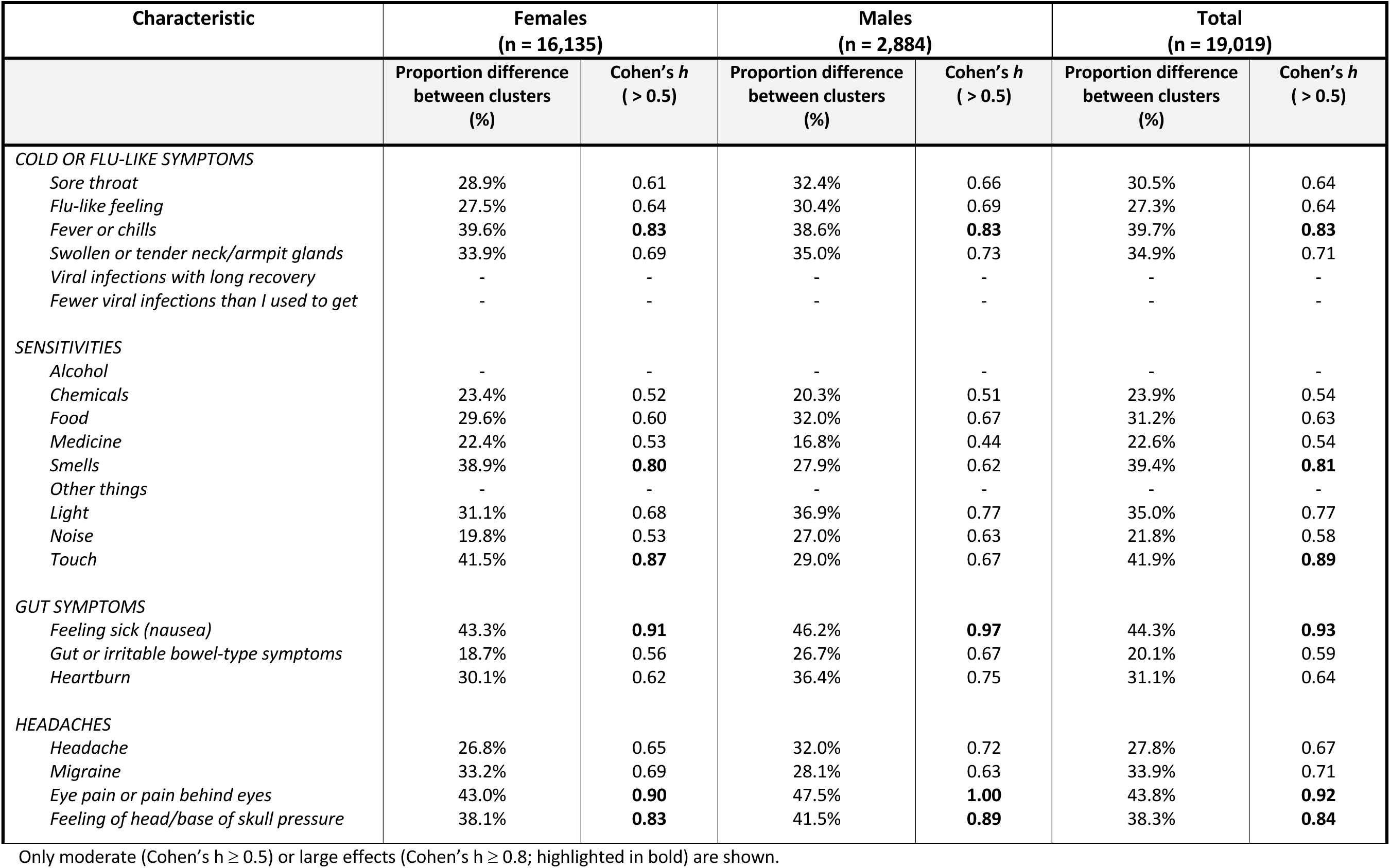
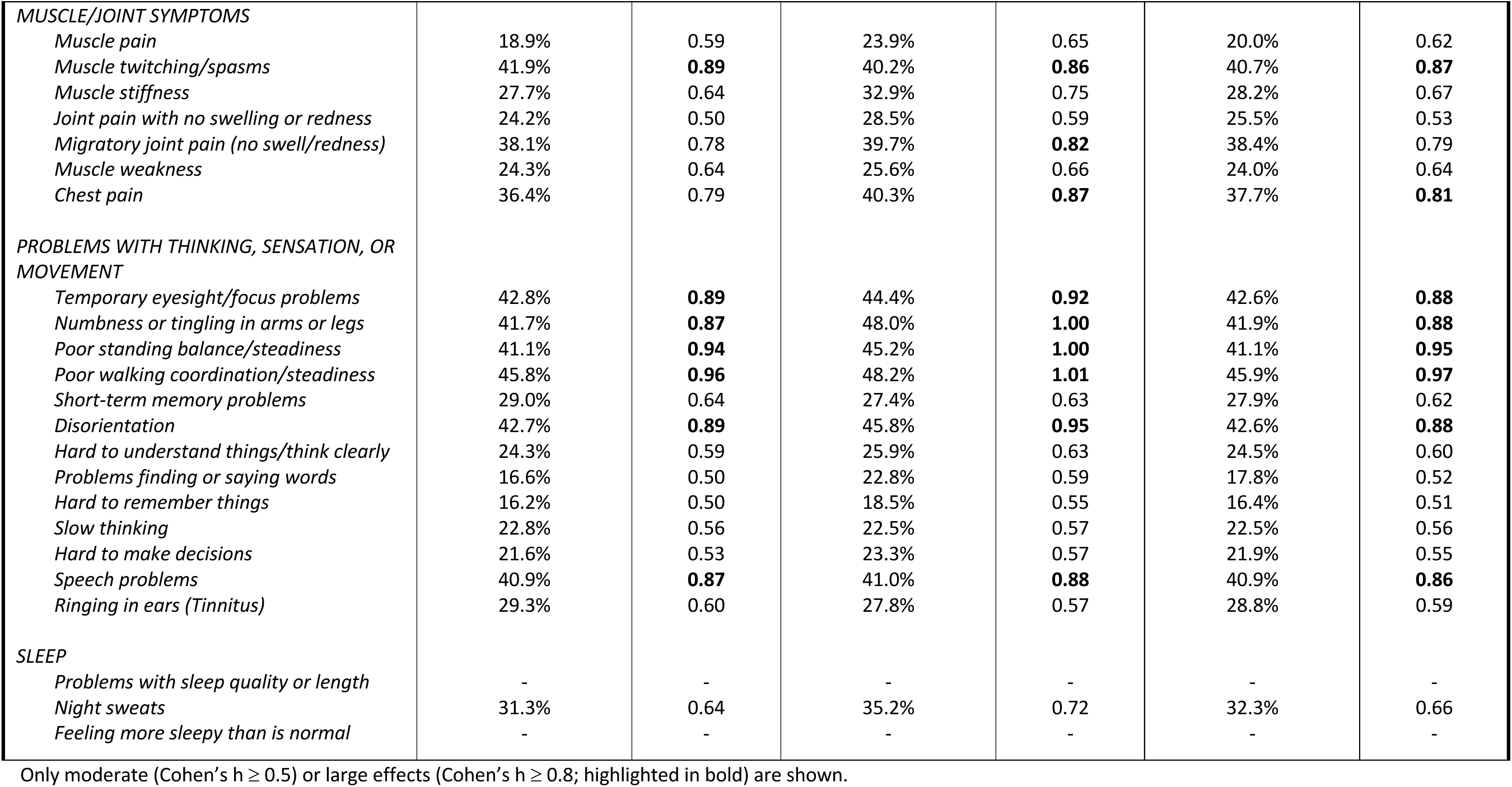
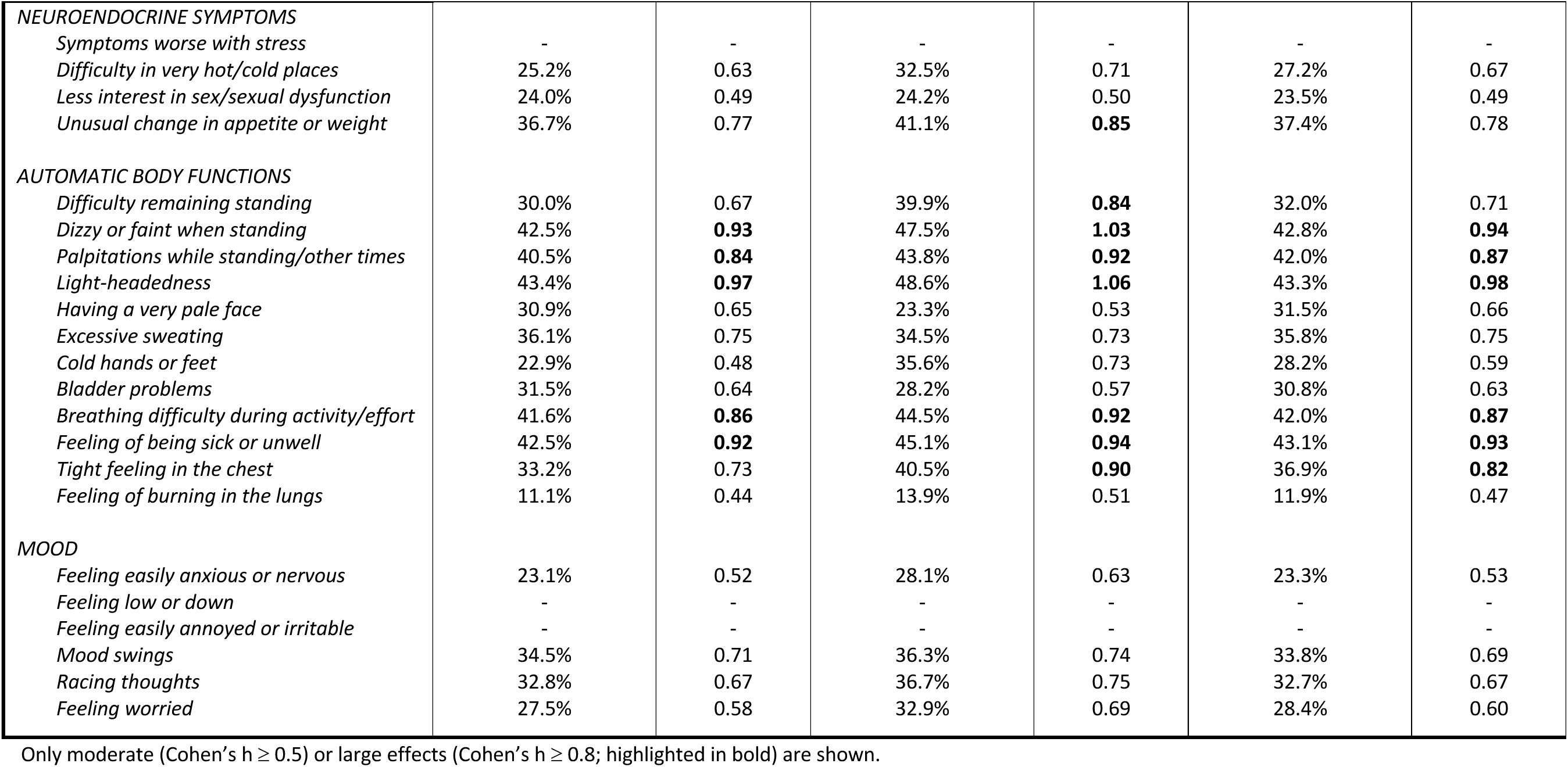
Influential symptoms driving cluster separation, based on Cohen’s h, for the total sample and sex-based subgroups.

### Inferential Analysis

Lastly, we explored the association between infectious disease onset and cluster membership (Table 2). In multivariable logistic regression adjusted for sex, age, deprivation, and ethnicity, infectious onset was associated with a 24% increased odds (OR = 1.24, 95% CI: 1.15–1.35), and unknown onset with 30% increased odds (OR = 1.30, 95% CI: 1.18–1.43) of belonging to the HSBC, relative to non-infectious disease onset. The association was similar for males and females, although estimation was imprecise for males due to reduced power.

**Table 2.** Unadjusted and adjusted odds ratios for the association between onset type and high symptom burden cluster membership for the total sample and sex-based subgroups.

|  | Females (n = 16,135) |  | Males (n = 2,884) |  | Total Sample (n = 19,019) |  |
| --- | --- | --- | --- | --- | --- | --- |
|  | OR [95% CI] | p-value | OR [95% CI] | p-value | OR [95% CI] | p-value |
| **UNADJUSTED** |  |  |  |  |  |  |
| *Onset Type* |  |  |  |  |  |  |
| Non-infectious | Reference | - | Reference | - | Reference | - |
| Infectious | 1.13 [1.04 – 1.25] | **2.00x10<sup>-03</sup>** | 1.16 [0.93 – 1.44] | 0.18 | 1.13 [1.04 – 1.22] | **3.53x10<sup>-03</sup>** |
| Unknown onset | 1.28 [1.15 – 1.41] | **2.82x10<sup>-06</sup>** | 1.20 [0.94– 1.54] | 0.14 | 1.24 [1.13 – 1.37] | **4.40x10<sup>-06</sup>** |
| **ADJUSTED** |  |  |  |  |  |  |
| *Onset Type* |  |  |  |  |  |  |
| Non-Infectious | Reference | - | Reference | - | Reference | - |
| Infectious | 1.25 [1.15 – 1.37] | **5.03x10<sup>-07</sup>** | 1.17 [0.94 – 1.46] | 0.15 | 1.24 [1.15 – 1.35] | **1.85x10<sup>-07</sup>** |
| Unknown | 1.33 [1.20 – 1.47] | **1.04x10<sup>-07</sup>** | 1.17 [0.91 – 1.50] | 0.23 | 1.30 [1.18 – 1.43] | **6.53x10<sup>-08</sup>** |
| *Sex* |  |  |  |  |  |  |
| Female |  |  |  |  | Reference | - |
| Male |  |  |  |  | 0.52 [0.48– 0.56] | **<2x10<sup>-16</sup>** |
| *Age* |  |  |  |  |  |  |
| 16 – 25 | Reference | - | Reference | - | Reference | - |
| 26 – 35 | 0.91 [0.76 – 1.07] | 0.26 | 1.46 [0.96 – 2.22] | 0.08 | 0.95 [0.81 – 1.11] | 0.54 |
| 36 – 45 | 0.88 [0.75 – 1.04] | 0.13 | 1.58 [1.07 – 2.36] | **0.02** | 0.93 [0.80 – 1.08] | 0.33 |
| 46 – 55 | 0.85 [0.73 – 1.00] | 0.05 | 1.64 [1.12 – 2.42] | **0.01** | 0.90 [0.78 – 1.04] | 0.15 |
| 56 – 65 | 0.62 [0.53 – 0.73] | **5.69x10<sup>-09</sup>** | 1.09 [0.74 – 1.61] | 0.66 | 0.65 [0.56 – 0.75] | **6.20x10<sup>-09</sup>** |
| 66 – 75 | 0.35 [0.29 – 0.41] | **<2x10<sup>-16</sup>** | 0.74 [0.49 – 1.11] | 0.14 | 0.37 [0.32 – 0.43] | **<2x10<sup>-16</sup>** |
| 76 and over | 0.21 [0.16 – 0.27] | **<2x10<sup>-16</sup>** | 0.42 [0.24 – 0.72] | **1.89x10<sup>-03</sup>** | 0.21 [0.17 – 0.26] | **<2x10<sup>-16</sup>** |
| *IMD Group* |  |  |  |  |  |  |
| Quintile 1 (most deprived) | Reference | - | Reference | - | Reference | - |
| Quintile 2 | 0.91 [0.82 – 1.00] | 0.05 | 0.83 [0.66 – 1.04] | 0.10 | 0.90 [0.82 – 0.98] | **0.01** |
| Quintile 3 | 0.87 [0.78 – 0.96] | **4.79x10<sup>-03</sup>** | 0.76 [0.61 – 0.96] | **0.02** | 0.86 [0.79 – 0.94] | **1.47x10<sup>-03</sup>** |
| Quintile 4 | 0.81 [0.73 – 0.89] | **3.52x10<sup>-05</sup>** | 0.88 [0.70 – 1.11] | 0.30 | 0.81 [0.74 – 0.89] | **7.33x10<sup>-06</sup>** |
| Quintile 5 (least deprived) | 0.74 [0.66 – 0.82] | **1.05x10<sup>-07</sup>** | 0.64 [0.49 – 0.83] | **8.89x10<sup>-04</sup>** | 0.69 [0.62 – 0.77] | **2.93x10<sup>-12</sup>** |
| *Ethnicity* |  |  |  |  |  |  |
| White | Reference | - | Reference | - | Reference | - |
| Ethnic Minority | 1.03 [0.85 – 1.25] | 0.73 | 1.50 [0.97 – 2.34] | 0.07 | 1.08 [0.91 – 1.29] | 0.37 |

An OR > 1 indicates a higher likelihood of High Symptom Burden Cluster membership. P-values < 0.05 are in bold. Ethnic minority = Asian, Asian British, Black, African, Caribbean, Black British, mixed or multiple ethnic minority groups, or other ethnic group. CI = confidence interval. IMD = index of multiple deprivation.

### Sex-Specific Clusters

A two-cluster solution was also found to be optimal in the subgroup analyses for both females (Supplemental Figures 3A-B, 4) and males (Supplemental Figures 3C-D, 5). Assessment of ARI values based on 100 bootstrap replicates showed the clusters for both subgroups were stable (female meanARI = 0.78, male meanARI = 0.91) (Supplemental Figure 6).

Clustering of females, and separately males, also each resulted in two clusters characterised as a HSBC and a LSBC (Supplemental Table 2). For females, a higher proportion of individuals were allocated to the HSBC (61.6%) compared to the LSBC (38.4%), whereas for males, the LSBC had a higher proportion of individuals (52.0%) compared to the HSBC (48.0%). Patterns in baseline characteristics across clusters for females and males are generally similar to the total sample, with differences in statistical significance likely due to the smaller number of males (n = 2,884) compared to females (n = 16,135). Symptom burden across both clusters was generally higher in females than males. There were also sex differences in the individual symptoms most strongly contributing to separation of the HSBC and LSBC (Table 1). Several symptoms within the pain, cognitive/neurological, autonomic, gut, and mood domains had larger between-cluster differences in males than females. After false discovery rate correction, only heartburn, extremity numbness or tingling, eye pain, and chest tightness showed statistically significant cluster-by-sex interactions; for each of these symptoms, the HSBC-LSBC contrast was larger in males than in females.

### Genome-Wide Association Study

No variant reached genome-wide significance at the standard threshold of p < 5 x 10^-8^. At a less stringent threshold of *p* < 8 x 10^-7^ three associations were identified (Figure 3, Supplemental Table 3), among which approximately one is expected to be a false positive discovery (20).

**Figure 3.**
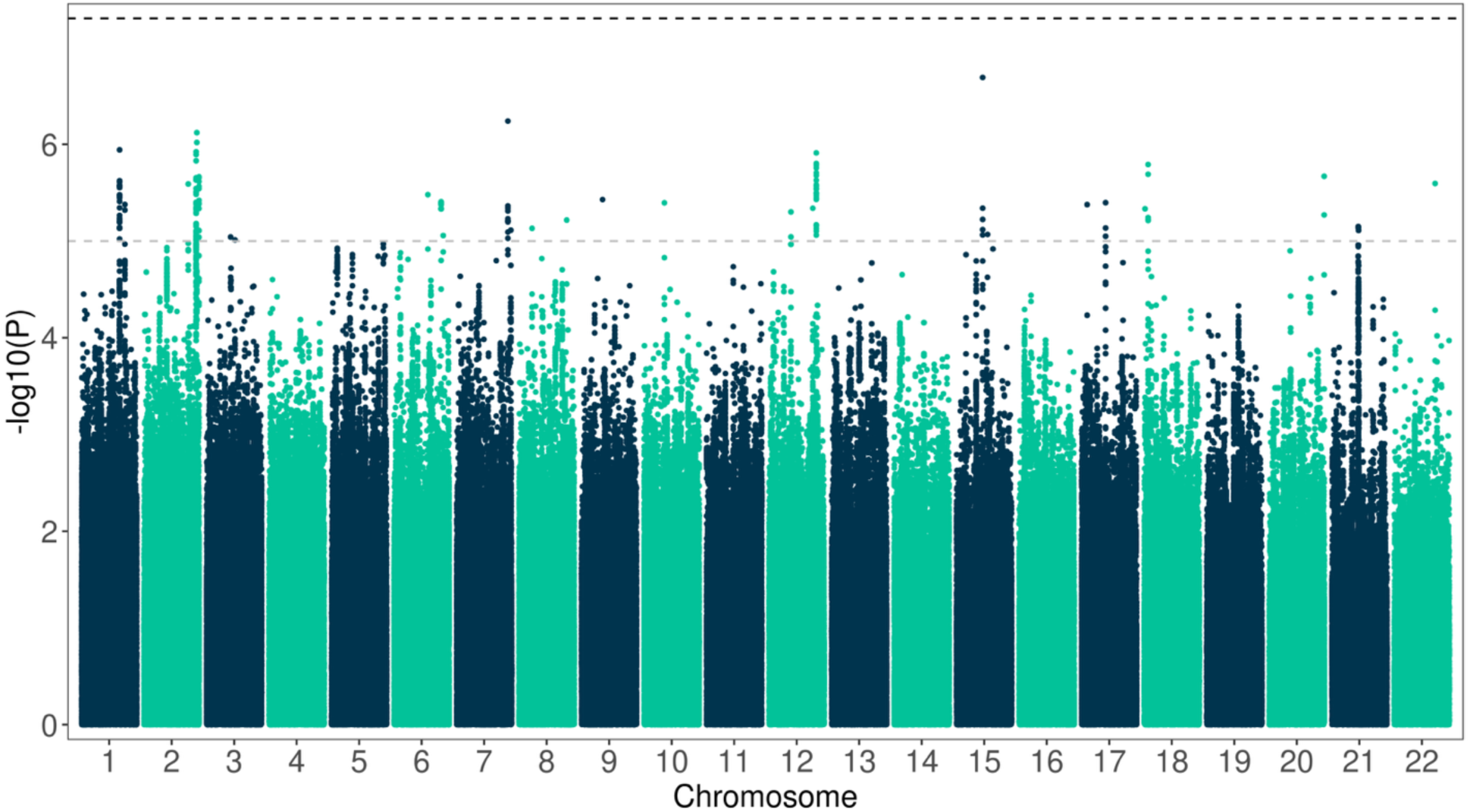
No genome-wide significant associations to HSBC/LSBC status. Manhattan plot showing the chromosomal position of DNA variants (x-axis) against −log10p (y-axis) from genome-wide association tests for HSBC status accounting for sex, genotyping batch and genetic ancestry. The upper horizontal dashed line indicates the threshold used to define genome-wide significant associations (*p* < 5 x 10^-8^); the lower horizontal dashed line indicates *p* < 1 x 10^-5^. Three loci on chromosome 15 (*RORA*), 7 (*RARRES2*) and 2 (*SPP2* / *TRPM8*) have associations with *p* < 8 x 10^-7^.

Nearby protein-coding genes are involved in chronobiology, inflammation and pain sensing (Supplemental Table 3).

## Discussion

This study applied a data-driven approach to identify symptom-based ME/CFS subtypes in the DecodeME cohort, using *k*-modes clustering and sex-stratified analyses. Two consistent clusters emerged: a HSBC and a LSBC. The HSBC was characterised by more symptoms, greater illness severity, greater proportion of people with worsening illness course, and higher comorbidity prevalence, particularly for fibromyalgia, IBS, and clinical depression. The HSBC included more females, younger individuals, and those from more deprived areas. Similar clustering patterns were observed in females and males, although females in the HSBC showed higher overall symptom prevalence and more pronounced sensory sensitivities, whereas males showed larger differences between clusters for cognitive, autonomic, and mood symptoms. Individuals with infectious or unknown onset were more likely to belong to the HSBC, even after accounting for differences in age, sex, ethnicity and socioeconomic deprivation. Although the identified genetic associations were suggestive rather than definitive, the proximity of the implicated loci to genes involved in chronobiology, inflammation, and pain processing is notable given the prominence of sleep disturbance, post-exertional malaise, and sensory hypersensitivity in ME/CFS.

Previous studies employing clustering techniques (e.g., latent class analysis, k-means, hierarchical agglomerative clustering) on ME/CFS symptom data from single and combined datasets revealed a polysymptomatic/high symptom burden cluster akin to the HSBC (21–29), an oligosymptomatic/low symptom burden cluster akin to the LSBC (22,24,25,28) and clusters characterised by a dominant symptom type, including pain (primarily muscle or joint) (37,39,42), PEM (27,28,31), mood symptoms (29,32,33), and autonomic dysfunction (e.g., GI symptoms, circulatory symptoms, dysautonomia) (26,32,34). Comparability between studies is limited by inconsistent or absent case definitions and heterogeneity in symptom lists and analytic inputs (e.g., raw vs factor-reduced data). Furthermore, sample sizes were frequently small (n < 160), and in some studies very small clusters (<10% of the sample) were reported, reducing statistical power, the reliability of rare subtypes, and external validity. Additionally, we identified only one study that conducted sex-stratified clustering (35), with very small male groups (<20 males per cluster), below recommended thresholds (53). Their findings aligned with our analyses, where females were more likely to be in the HSBC and exhibited higher symptom prevalence overall.

The observed association between infectious or unknown onset and membership in the HSBC suggests that these onset types could delineate clinically and potentially biologically distinct ME/CFS subtypes. Individuals with an infectious onset were more likely to experience severe symptomatology, supporting prior evidence that post-infectious cases can result in a more debilitating disease course (25). This aligns with longitudinal findings from post-viral fatigue studies, such as those involving Epstein-Barr virus and COVID-19, where the severity of the initial illness predicted long-term symptom burden (25,36).

Interestingly, individuals who reported not knowing whether an infection was involved at onset also showed increased odds of being in the HSBC. However, interpretation is limited by the survey wording: participants were asked only whether an infection occurred ‘when, or just before, [their] first ME/CFS symptoms started’, not about other possible triggers or whether the onset circumstances were uncertain. Consequently, the ‘don’t know’ response may conflate several distinct scenarios, including uncertainty about infection involvement, genuinely unknown onset circumstances, and non-infectious triggering events. Future studies could improve onset characterization by distinguishing between infection-related onset, specific non-infectious triggers, no identifiable trigger, and lack of information about onset.

These clinical patterns can be considered alongside genetic findings from DecodeME, while distinguishing between ME/CFS risk loci and the exploratory GWAS undertaken in the present subtype analysis. The wider DecodeME GWAS identified eight genome-wide significant loci associated with ME/CFS risk, including loci near genes involved in immune response and neurological function, and reported stronger genetic signals among participants with infection at onset (4). This provides broader support for immunological and neurological contributions to ME/CFS and is consistent with our observation that infectious onset was associated with membership of the HSBC. In contrast, the GWAS conducted as part of the present analysis did not identify any genome-wide significant subtype-associated variants, and the three suggestive associations should therefore be interpreted cautiously, particularly as approximately one would be expected to be false positive at the threshold used. Nevertheless, the proximity of these suggestive loci to genes implicated in circadian regulation, inflammatory signalling, and sensory or pain processing is notable in relation to the clinical profile of the HSBC, which was marked by greater illness severity, sleep-related symptoms, sensory sensitivities, pain-associated comorbidities, and higher prevalence of infectious or uncertain onset. Taken together, the genetic findings do not validate the symptom clusters directly, but they provide a plausible biological frame for interpreting them and suggest that symptom burden, onset type, sex, and comorbidity profile may be important stratification variables in future genetic and mechanistic studies of ME/CFS.

The primary strength of this study is its large sample size, which enabled sex-stratified clustering and analysis of raw symptom data, capturing a wide range of symptoms without pre-defined assumptions. Despite the cohort being predominantly White, it included more ethnic minority participants than previous UK ME/CFS studies, enabling preliminary exploration of cluster distribution across ethnic groups. Additional strengths include requiring a clinical diagnosis and applying both clinical and research case definitions, ensuring a heterogeneous yet relevant sample. The *k*-modes clustering method was internally validated and showed high cluster stability, even in the smaller male subgroup.

This study has several limitations. First, there is the potential for Type 1 errors given the large sample size and extensive number of variables, although corrections for multiple testing were applied to reduce this risk. Additionally, clustering may reflect sample-specific patterns despite internal validation. To address this, future studies should aim to externally validate the clusters using independent ME/CFS datasets with similar symptom variables to assess replicability and generalisability. Meta-clustering approaches that integrate results across multiple datasets or reapplying the clustering solution to new cohorts could further strengthen confidence in the identified symptom subgroups. Second, self-selection bias is another potential limitation.

Recruitment via online platforms and patient networks may have resulted in an overrepresentation of individuals who are more health literate, motivated, or able to participate. This may bias the sample towards individuals with particular demographic or illness characteristics. For instance, the mental and physical burden of participation could deter those with more severe ME/CFS, potentially leading to underestimation of the effect sizes observed, particularly those relating to symptom severity or comorbidity burden. Older individuals and people from ethnic minority groups appear underrepresented in ME/CFS cohorts, consistent with English GP data showing fewer diagnoses in older individuals than expected and disproportionate underdiagnosis among ethnic minority groups (37), as well as underrepresentation of older individuals in DecodeME (4). The results of this study are also not generalisable to paediatric or adolescent populations, whose presentation of ME/CFS can differ from that of adults (38).

While the clustering analysis identifies symptom-based subgroups, these groupings may still have relevance for clinical care. Because current ME/CFS management guidance and care recommendations include severity-based adaptations (39), healthcare professionals likely already respond differently to patients with high versus low symptom burden; cluster characteristics could therefore support a more structured understanding of patient heterogeneity. This may help guide awareness of differential needs, particularly in settings where a more nuanced approach is not consistently applied. For example, individuals in the high symptom burden group may benefit from more proactive care or support, including signposting to services such as social care, occupational therapy, or benefit systems to help manage functional limitations. These findings emphasise the importance of recognising heterogeneity in ME/CFS and underscore the need to address the specific presentation of the individual, rather than relying on a uniform model of the disease.

The association between infectious or unknown onset and membership in the HSBC membership underscores the value of incorporating onset type into clinical assessments. Although most individuals recover fully from infections, a subset develop ME/CFS (40), and among those with ME/CFS, individuals with an infectious onset are more likely to carry a higher symptom burden. Closer monitoring following certain infections, particularly in cases involving prolonged or atypical recovery, may help identify early signs of ME/CFS. However, given the low absolute risk of ME/CFS following most infections (40), any targeted post-infection surveillance strategies must be proportionate and guided by further research. These findings also highlight the importance of mechanistic studies to better understand why infectious onset may be associated with more severe disease, potentially revealing biological pathways that could inform both preventive strategies and therapeutic development.

Although curative treatment remains the ideal goal, no effective interventions currently exist for ME/CFS (41). In the meantime, multidisciplinary care focused on symptom management is essential. This study reinforces the importance of ensuring that such care is widely accessible and responsive to complex clinical needs. Symptom management may include advice on pacing, comorbidity screening, sensory modulation, social and psychological support, and pharmacologic treatment to alleviate specific symptoms (42).

While nearly all symptoms were less prevalent in the LSBC, reflecting a generally milder and less multisystem pattern, this was not an inevitable outcome given the known heterogeneity of ME/CFS symptoms. That such a distinction emerged underscores the potential value of clustering approaches in elucidating population-level symptom patterns. We selected k-modes because it is well suited to large categorical datasets and was found to perform well in a recent study using a similar type of dataset (14). Work is also needed to determine whether clusters correspond to objective clinical or physiological measures, predict meaningful outcomes, and remain stable over time. In parallel, combining genetic data from studies like DecodeME with detailed clinical features, such as symptom-based clusters and onset history, may further refine ME/CFS subtypes and help reveal underlying biological pathways, supporting the development of more precise diagnostic tools and targeted interventions.

## Conclusions

This clustering analysis of ME/CFS symptoms in the DecodeME study applied an unsupervised machine learning approach to the largest cohort to date, identifying two symptom-based clusters. These findings reinforce the heterogeneity of ME/CFS and are consistent with prior research, while benefiting from greater statistical power and demographic breadth. The observed associations between symptom burden, sex, and onset type, and particularly the heightened symptom severity linked to infectious and unknown onset, suggest the existence of clinically meaningful subtypes, potentially reflecting distinct biological mechanisms.

## Data Availability

Due to privacy and ethical considerations, the data are not publicly available. Access to DecodeME can be granted to bona fide researchers and will require approval from the DecodeME Data Access Committee.

https://institute-genetics-cancer.ed.ac.uk/decodeme-the-worlds-largest-mecfs-study/researcher-access

## Acknowledgements

The authors would like to thank the DecodeME participants and DecodeME staff for their contributions to this study. The authors would like to acknowledge that DecodeME was funded by the National Institute for Health and Care Research (NIHR) and Medical Research Council (MRC), grant number MC_PC_20005, and that JJD’s research is funded by generous donations. The authors would like to thank Sian Leary for providing thoughtful feedback on this preprint and Lauren DeLong for providing early guidance on cluster analysis. The authors would also like to acknowledge that this work made use of the resources provided by the Edinburgh Compute and Data Facility.

