## Supplemental for "PREPRINT: Cluster analysis of ME/CFS symptoms in DecodeME reveals two subgroups and a link to onset type"

### Supplemental Information

**Supplemental Table 1**. Baseline characteristics of the study population.

| Characteristic | N (%) |
|---|---|
| ***Sex, female*** | 16,135 (84.8%) |
| ***Age (mean, SD) - years*** | 48.7 (14.5) |
| ***Ethnic Group***+ | |
| *Ethnic minority* | 590 (3.1%) |
| *White* | 18429 (96.9%) |
| ***IMD Group*** | |
| *Quintile 1 (most deprived)* | 4036 (21.2%) |
| *Quintile 2* | 4639 (24.4%) |
| *Quintile 3* | 3972 (20.9%) |
| *Quintile 4* | 3856 (20.3%) |
| *Quintile 5 (least deprived)* | 2516 (13.2%) |
| ***Onset Type*** | |
| *Non-infectious* | 3119 (16.4%) |
| *Infectious* | 11628 (61.1%) |
| *Unknown* | 4272 (22.5%) |
| ***Length of Illness*** | |
| *6 months to 1 year* | 288 (1.5%) |
| *1 – 3 years* | 1675 (8.8%) |
| *3 – 5 years* | 1936 (10.2%) |
| *5 – 10 years* | 3909 (20.6%) |
| *10 years and over* | 11211 (58.9%) |
| ***Illness Severity*** | |
| *Mild* | 5333 (28.1%) |
| *Moderate* | 11053 (58.1%) |
| *Severe* | 2475 (13.0%) |
| *Very Severe* | 158 (0.8%) |
| ***Illness Course*** | |
| *Getting worse* | 3393 (17.8%) |
| *Relapsing and remitting* | 2136 (11.2%) |
| *Fluctuating* | 11457 (60.2%) |
| *No change* | 1545 (8.1%) |
| *Getting better* | 475 (2.5%) |
| *Recovered* | 13 (0.01%) |
| ***Comorbidities*** | |
| *Adrenal insufficiency* | 297 (1.6%) |
| *Mast cell activation syndrome* | 519 (2.7%) |
| *Hepatitis* | 252 (1.3%) |
| *HIV/AIDS* | 95 (0.5%) |
| *Irritable bowel syndrome* | 8311 (43.7%) |
| *Lupus* | 167 (0.9%) |
| *Lyme disease* | 413 (2.2%) |
| *Clinical depression* | 6915 (36.4%) |
| *Multiple sclerosis* | 113 (0.6%) |
| *Myasthenia gravis* | 95 (0.5%) |
| *Narcolepsy* | 102 (0.5%) |

* SD = standard deviation, + ethnic minority = Asian, Asian British, Black, African, Caribbean, Black British, mixed or multiple ethnic groups, or other ethnic minority group, IMD = index of multiple deprivation

| Characteristic | N (%) |
|---|---|
| ***Comorbidities*** | |
| *Anemia* | 2965 (15.6%) |
| *Overactive adrenal glands* | 94 (0.5%) |
| *Overactive thyroid* | 424 (2.2%) |
| *Parkinson's disease* | 81 (0.4%) |
| *Polymyositis* | 79 (0.4%) |
| *Rheumatoid arthritis* | 604 (3.2%) |
| *Schizophrenia* | 101 (0.5%) |
| *Sleep Apnoea* | 1097 (5.8%) |
| *Tuberculosis* | 123 (0.7%) |
| *Underactive thyroid* | 2603 (13.7%) |
| *B12 deficiency not treatable* | 1514 (8.0%) |
| *Polymyalgia rheumatica* | 211 (1.1%) |
| *Fibromyalgia* | 6621 (34.8%) |
| *Sarcoidosis* | 134 (0.7%) |
| *Shingles* | 1790 (9.4%) |
| *Sjogren's syndrome* | 317 (1.7%) |
| *Upper airway resistance syndrome* | 102 (0.5%) |
| *Q fever* | 295 (1.6%) |
| *Bipolar disorder* | 394 (2.1%) |
| *Cancer* | 936 (4.9%) |
| *Coeliac disease* | 584 (3.1%) |
| *Diabetes* | 1224 (6.4%) |
| *Haemochromatosis* | 187 (1.0%) |
| ***SYMPTOMS IN THE PAST 6 MONTHS*** | |
| *COLD OR FLU-LIKE SYMPTOMS* | |
| *Sore throat* | 11919 (62.7%) |
| *Flu-like feeling* | 14291 (75.1%) |
| *Fever or chills* | 8373 (44.0%) |
| *Swollen or tender neck/armpit glands* | 10547 (55.5%) |
| *Viral infections with long recovery* | 7488 (39.4%) |
| *Fewer viral infections than before* | 2108 (11.1%) |
| *SENSITIVITIES* | |
| *Alcohol* | 8938 (47.0%) |
| *Chemicals* | 5929 (31.2%) |
| *Food* | 9696 (51.0%) |
| *Medicine* | 4961 (26.1%) |
| *Smells* | 9591 (50.4%) |
| *Other things* | 3764 (19.8%) |
| *Light* | 13056 (68.7%) |
| *Noise* | 15649 (82.3%) |
| *Touch* | 8274 (43.5%) |
| *HEADACHES* | |
| *Headache* | 14684 (77.2%) |
| *Migraine* | 7861 (41.3%) |
| *Eye pain or pain behind eyes* | 11500 (60.5%) |
| *Feeling of head/base of skull pressure* | 13019 (68.5%) |

| **Characteristic** | **N (%)** |
|---|---|
| ***SYMPTOMS IN THE PAST 6 MONTHS*** | |
| *GUT SYMPTOMS* | |
| *Feeling sick (nausea)* | 11457 (60.2%) |
| *Gut or irritable bowel-type symptoms* | 16392 (86.2%) |
| *Heartburn* | 8809 (46.3%) |
| *MUSCLE/JOINT SYMPTOMS* | |
| *Muscle pain* | 16594 (87.3%) |
| *Muscle twitching/spasms* | 12282 (64.6%) |
| *Muscle stiffness* | 14499 (76.2%) |
| *Joint pain with no swelling or redness* | 12019 (63.2%) |
| *Migratory joint pain (no swell/no red)* | 10713 (56.3%) |
| *Muscle weakness* | 15677 (82.4%) |
| *Chest pain* | 7393 (38.9%) |
| *PROBLEMS WITH THINKING, SENSATION, OR MOVEMENT* | |
| *Temporary eyesight/focus problems* | 10076 (53.0%) |
| *Numbness or tingling in arms or legs* | 11089 (58.3%) |
| *Poor standing balance/steadiness* | 13716 (72.1%) |
| *Poor walking coordination/steadiness* | 11551 (60.7%) |
| *Confusion or 'brainfog'* | 18047 (94.9%) |
| *Finding it hard to concentrate* | 18060 (95.0%) |
| *Short-term memory problems* | 13618 (71.6%) |
| *Disorientation* | 9310 (49.0%) |
| *Hard to understand things/think clearly* | 14976 (78.7%) |
| *Problems finding or saying words* | 16430 (86.4%) |
| *Hard to remember things* | 16689 (87.8%) |
| *Slow thinking* | 15113 (79.5%) |
| *Hard to make decisions* | 15147 (79.6%) |
| *Speech problems* | 8234 (43.3%) |
| *Ringing in ears (Tinnitus)* | 10195 (53.6%) |
| *SLEEP* | |
| *Problems with sleep quality or length* | 15978 (84.0%) |
| *Night sweats* | 10229 (53.8%) |
| *Feeling more sleepy than is normal* | 15064 (79.2%) |
| *Unrefreshing sleep* | 18488 (97.2%) |
| *NEUROENDOCRINE SYMPTOMS* | |
| *Symptoms worse with stress* | 16208 (85.2%) |
| *Difficulty coping with being in very hot or cold places* | 14888 (78.3%) |
| *Less interest in sex and/or sexual function difficulties* | 11734 (61.7%) |
| *Unusual change in appetite or weight* | 11719 (61.6%) |

| Characteristic | N (%) |
|---|---|
| ***SYMPTOMS IN THE PAST 6 MONTHS*** | |
| *AUTOMATIC BODY FUNCTIONS* | |
| *Difficulty remaining standing* | 13319 (70.0%) |
| *Dizzy or faint when standing* | 12819 (67.4%) |
| *Palpitations while standing/other times* | 10583 (55.6%) |
| *Light-headedness* | 13299 (69.9%) |
| *Having a very pale face* | 7728 (40.6%) |
| *Excessive sweating* | 8346 (43.9%) |
| *Cold hands or feet* | 12237 (64.3%) |
| *Bladder problems* | 10977 (57.7%) |
| *Breathing difficulty during activity/effort* | 10198 (53.6%) |
| *Feeling of being sick or unwell* | 12480 (65.6%) |
| *Tight feeling in the chest* | 6794 (35.7%) |
| *Feeling of burning in the lungs* | 1755 (9.2%) |
| *MOOD* | |
| *Feeling easily anxious or nervous* | 13917 (73.2%) |
| *Feeling low or down* | 15535 (81.7%) |
| *Feeling easily annoyed or irritable* | 14252 (74.9%) |
| *Mood swings* | 9273 (48.8%) |
| *Racing thoughts* | 9854 (51.9%) |
| *Feeling worried* | 12240 (64.4%) |
| *FATIGUE* | |
| *Do you have fatigue (lack of energy) often, repeatedly or all the time?* | 19019 (100%) |
| *I feel like a battery that can never fully recharge, even when I rest* | 18910 (99.4%) |
| *I feel both physically and mentally fatigued* | 18656 (98.1%) |
| *I often feel fatigue, and this can get worse when I'm active* | 19019 (100%) |
| *I have this fatigue (lack of energy) more than half the time* | 17946 (94.4%) |
| *My fatigue (lack of energy) is disabling* | 17701 (93.1%) |
| *POST-EXERTIONAL MALAISE* | |
| *The change in my symptoms lasts a long time, which can be 24 hours or more* | 19019 (100%) |
| *My symptoms (such as pain, fatigue, or feeling out-of-sorts) get worse, or I get new symptoms, and this reduces how much I can do* | 19019 (100%) |

**Supplemental Table 2.** Baseline characteristics for the total sample and sex-based subgroups, stratified by cluster.

| Characteristic | Females (n = 16,135) | | | Males (n = 2,884) | | | Total (n = 19,019) | | |
|---|---|---|---|---|---|---|---|---|---|
| | HSBC (n = 9,932, 61.6%) | LSBC (n = 6,203, 38.4%) | p-value | HSBC (n = 1,385, 48.0%) | LSBC (n = 1,499, 52.0%) | p-value | HSBC n = 10,849 (57.0%) | LSBC (n = 8,170, 43.0%) | p-value |
| ***Sex, female (n, %)*** | - | - | | - | - | | 9629 (88.8%) | 6506 (79.6%) | **<0.001** |
| ***Age (mean, SD)*** | 46.6 (13.6) | 51.1 (15.2) | **<0.001** | 49.2 (13.6) | 52.9 (15.3) | **<0.001** | 46.7 (13.6) | 51.5 (15.2) | **<0.001** |
| ***Ethnic Group⁺ (n, %)*** | | | 0.98 | | | 1.00 | | | 0.2 |
| *Ethnic minority* | 324 (3.3%) | 176 (2.8%) | | 55 (4.0%) | 35 (2.3%) | | 364 (3.4%) | 226 (2.8%) | |
| *White* | 9608 (96.7%) | 6027 (97.2%) | | 1330 (96.0%) | 1464 (97.7%) | | 10485 (96.6%) | 7944 (97.2%) | |
| ***IMD Group (n, %)*** | | | **<0.001** | | | 0.05 | | | **<0.001** |
| *Quintile 1 (most deprived)* | 2246 (22.6%) | 1200 (19.3%) | | 314 (22.7%) | 276 (18.4%) | | 2466 (22.7%) | 1570 (19.2%) | |
| *Quintile 2* | 2472 (24.9%) | 1471 (23.7%) | | 334 (24.1%) | 362 (24.1%) | | 2696 (24.9%) | 1943 (23.8%) | |
| *Quintile 3* | 2061 (20.8%) | 1297 (20.9%) | | 278 (20.1%) | 336 (22.4%) | | 2261 (20.8%) | 1711 (20.9%) | |
| *Quintile 4* | 1941 (19.5%) | 1320 (21.3%) | | 295 (21.3%) | 300 (20.0%) | | 2134 (19.7%) | 1722 (21.1%) | |
| *Quintile 5 (least deprived)* | 1212 (12.2%) | 915 (14.8%) | | 164 (11.8%) | 225 (15.0%) | | 1292 (11.9%) | 1224 (15.0%) | |
| ***Onset Type (n, %)*** | | | **<0.001** | | | 1.00 | | | **<0.001** |
| *Non-infectious* | 1576 (15.9%) | 1129 (18.2%) | | 185 (13.4%) | 229 (15.3%) | | 1686 (15.5%) | 1433 (17.5%) | |
| *Infectious* | 6034 (60.8%) | 3771 (60.8%) | | 881 (63.6%) | 942 (62.8%) | | 6625 (61.1%) | 5003 (61.2%) | |
| *Unknown* | 2322 (23.4%) | 1303 (21.0%) | | 319 (23.0%) | 328 (21.9%) | | 2538 (23.4%) | 1734 (21.2%) | |
| ***Length of Illness (n, %)*** | | | 0.07 | | | 1.00 | | | 0.60 |
| *6 months to 1 year* | 142 (1.4%) | 111 (1.8%) | | 23 (1.7%) | 12 (0.8%) | | 155 (1.4%) | 133 (1.6%) | |
| *1 – 3 years* | 880 (8.9%) | 563 (9.1%) | | 110 (7.9%) | 122 (8.1%) | | 948 (8.7%) | 727 (8.9%) | |
| *3 – 5 years* | 1029 (10.4%) | 665 (10.7%) | | 109 (7.9%) | 133 (8.9%) | | 1102 (10.2%) | 834 (10.2%) | |
| *5 – 10 years* | 2129 (21.4%) | 1200 (21.4%) | | 282 (20.4%) | 298 (19.9%) | | 2309 (21.3%) | 1600 (19.6%) | |
| *10 years and over* | 5752 (57.9%) | 3664 (59.1%) | | 861 (62.2%) | 934 (62.3%) | | 6335 (58.4%) | 4876 (59.7%) | |
| ***Illness Severity (n, %)*** | | | **<0.001** | | | **<0.001** | | | **<0.001** |
| *Mild* | 2102 (21.2%) | 2443 (39.4%) | | 249 (18.0%) | 539 (36.0%) | | 2238 (20.6%) | 3095 (37.9%) | |
| *Moderate* | 5942 (59.8%) | 3387 (54.6%) | | 872 (63.0%) | 852 (56.8%) | | 6512 (60.0%) | 4541 (55.6%) | |
| *Severe* | 1774 (17.9%) | 356 (5.7%) | | 247 (17.8%) | 98 (6.5%) | | 1973 (18.2%) | 502 (6.1%) | |
| *Very Severe* | 114 (1.1%) | 17 (0.3%) | | 17 (1.2%) | 10 (0.7%) | | 126 (1.2%) | 32 (0.4%) | |

* SD = standard deviation, ⁺ ethnic minority = Asian, Asian British, Black, African, Caribbean, Black British, mixed or multiple ethnic groups, or other ethnic minority group, HSBC = high symptom burden cluster, IMD = index of multiple deprivation, LSBC = lower symptom burden cluster. P-values < 0.05 are shown in bold.

| Characteristic | Females (n = 16,135) | | | Males (n = 2,884) | | | Total (n = 19,019) | | |
|---|---|---|---|---|---|---|---|---|---|
| | HSBC (n = 9,932, 61.6%) | LSBC (n = 6,203, 38.4%) | p-value | HSBC (n = 1,385, 48.0%) | LSBC (n = 1,499, 52.0%) | p-value | HSBC (n = 10,849 57.0%) | LSBC (n = 8,170, 43.0%) | p-value |
| ***Illness Course (n, %)*** | | | **<0.001** | | | **<0.001** | | | **<0.001** |
| *Getting worse* | 2193 (22.1%) | 640 (10.3%) | | 333 (24.0%) | 227 (15.1%) | | 2463 (22.7%) | 930 (11.4%) | |
| *Relapsing and remitting* | 901 (9.1%) | 914 (14.7%) | | 138 (10.0%) | 183 (12.2%) | | 978 (9.0%) | 1158 (14.2%) | |
| *Fluctuating* | 6050 (60.9%) | 3844 (62.0%) | | 758 (54.7%) | 805 (53.7%) | | 6512 (60.0%) | 4945 (60.5%) | |
| *No change* | 667 (6.7%) | 529 (8.5%) | | 137 (9.9%) | 212 (14.1%) | | 770 (7.1%) | 775 (9.5%) | |
| *Getting better* | 121 (1.2%) | 268 (4.3%) | | 19 (1.4%) | 67 (4.5%) | | 126 (1.2%) | 349 (4.3%) | |
| *Recovered* | 0 | 8 (0.1%) | | 0 | 5 (0.3%) | | 0 | 13 (0.2%) | |
| ***Comorbidities (n, %)*** | | | | | | | | | |
| *Adrenal insufficiency* | 179 (1.8%) | 75 (1.2%) | 0.14 | 24 (1.7%) | 19 (1.3%) | 1.00 | 201 (1.9%) | 96 (1.2%) | **<0.001** |
| *Mast cell activation syndrome* | 383 (3.9%) | 83 (1.3%) | **<0.001** | 38 (2.7%) | 15 (1.0%) | **<0.001** | 411 (3.8%) | 108 (1.3%) | **<0.001** |
| *Hepatitis* | 122 (1.2%) | 84 (1.4%) | 1.00 | 24 (1.7%) | 22 (1.5%) | 1.00 | 139 (1.3%) | 113 (1.4%) | 1.00 |
| *HIV/AIDS* | 41 (0.4%) | 27 (0.4%) | 1.00 | 15 (1.1%) | 12 (0.8%) | 1.00 | 56 (0.5%) | 39 (0.5%) | 1.00 |
| *Irritable bowel syndrome* | 5257 (52.9%) | 2054 (33.1%) | **<0.001** | 643 (46.4%) | 357 (23.8%) | **<0.001** | 5693 (52.5%) | 2618 (32.0%) | **<0.001** |
| *Lupus* | 101 (1.0%) | 47 (0.8%) | 1.00 | 13 (0.9%) | 6 (0.4%) | 1.00 | 114 (1.1%) | 53 (0.6%) | 0.14 |
| *Lyme disease* | 238 (2.4%) | 105 (1.7%) | 0.11 | 41 (3.0%) | 29 (1.9%) | 1.00 | 273 (2.5%) | 140 (1.7%) | **<0.001** |
| *Clinical depression* | 4225 (42.5%) | 1846 (29.8%) | **<0.001** | 485 (35.0%) | 359 (23.9%) | **<0.001** | 4544 (41.9%) | 2371 (29.0%) | **<0.001** |
| *Multiple sclerosis* | 64 (0.6%) | 36 (0.6%) | 1.00 | 7 (0.5%) | 6 (0.4%) | 1.00 | 70 (0.6%) | 43 (0.5%) | 1.00 |
| *Myasthenia gravis* | 49 (0.5%) | 32 (0.5%) | 1.00 | 7 (0.5%) | 7 (0.5%) | 1.00 | 56 (0.5%) | 39 (0.5%) | 1.00 |
| *Narcolepsy* | 56 (0.6%) | 29 (0.5%) | 1.00 | 12 (0.9%) | 5 (0.3%) | 0.10 | 67 (0.6%) | 35 (0.4%) | 1.00 |
| *Anemia* | 2028 (20.4%) | 837 (13.5%) | **<0.001** | 66 (4.8%) | 34 (2.3%) | **0.01** | 2051 (18.9%) | 914 (11.2%) | **<0.001** |
| *Overactive adrenal glands* | 52 (0.5%) | 28 (0.5%) | 1.00 | 8 (0.6%) | 6 (0.4%) | 1.00 | 59 (0.5%) | 35 (0.4%) | 1.00 |
| *Overactive thyroid* | 249 (2.5%) | 137 (2.2%) | 1.00 | 22 (1.6%) | 16 (1.1%) | 1.00 | 264 (2.4%) | 160 (2.0%) | 1.00 |
| *Parkinson's disease* | 41 (0.4%) | 27 (0.4%) | 1.00 | 7 (0.5%) | 6 (0.4%) | 1.00 | 47 (0.4%) | 34 (0.4%) | 1.00 |
| *Polymyositis* | 41 (0.4%) | 27 (0.4%) | 1.00 | 5 (0.4%) | 6 (0.4%) | 1.00 | 46 (0.4%) | 33 (0.4%) | 1.00 |
| *Rheumatoid arthritis* | 377 (3.8%) | 146 (2.4%) | **<0.001** | 56 (4.0%) | 25 (1.7%) | **<0.001** | 417 (3.8%) | 187 (2.3%) | **<0.001** |
| *Schizophrenia* | 54 (0.5%) | 31 (0.5%) | 1.00 | 11 (0.8%) | 5 (0.3%) | 1.00 | 65 (0.6%) | 36 (0.4%) | 1.00 |
| *Sleep Apnoea* | 575 (5.8%) | 231 (3.7%) | **<0.001** | 172 (12.4%) | 119 (7.9%) | **<0.001** | 718 (6.6%) | 379 (4.6%) | **<0.001** |
| *Tuberculosis* | 71 (0.7%) | 38 (0.6%) | 1.00 | 6 (0.4%) | 8 (0.5%) | 1.00 | 75 (0.7%) | 48 (0.6%) | 1.00 |
| *Underactive thyroid* | 1507 (15.2%) | 903 (14.6%) | 1.00 | 92 (6.6%) | 101 (6.7%) | 1.00 | 1546 (14.3%) | 1057 (12.9%) | 0.33 |
| *B12 deficiency not treatable* | 1044 (10.5%) | 339 (5.5%) | **<0.001** | 86 (6.2%) | 45 (3.0%) | **<0.001** | 1108 (10.2%) | 406 (5.0%) | **<0.001** |
| *Polymyalgia rheumatica* | 115 (1.2%) | 71 (1.1%) | 1.00 | 14 (1.0%) | 11 (0.7%) | 1.00 | 125 (1.2%) | 86 (1.1%) | 1.00 |
| *Fibromyalgia* | 4638 (46.7%) | 1412 (22.8%) | **<0.001** | 405 (29.2%) | 166 (11.1%) | **<0.001** | 4902 (45.2%) | 1719 (21.0%) | **<0.001** |
| *Sarcoidosis* | 65 (0.7%) | 40 (0.6%) | 1.00 | 11 (0.8%) | 18 (1.2%) | 1.00 | 75 (0.7%) | 59 (0.7%) | 1.00 |
| *Shingles* | 1030 (10.4%) | 536 (8.6%) | **0.01** | 116 (8.3%) | 109 (7.3%) | 1.00 | 1105 (10.2%) | 685 (8.4%) | **<0.001** |
| *Sjogren's syndrome* | 217 (2.2%) | 74 (1.2%) | **<0.001** | 15 (1.1%) | 11 (0.7%) | 1.00 | 228 (2.1%) | 89 (1.1%) | **<0.001** |

HSBC = high symptom burden cluster, LSBC = lower symptom burden cluster. P-values < 0.05 are shown in bold.

| Characteristic | Females (n = 16,135) | | | Males (n = 2,884) | | | Total (n = 19,019) | | |
|---|---|---|---|---|---|---|---|---|---|
| | HSBC (n = 9,932, 61.6%) | LSBC (n = 6,203, 38.4%) | p-value | HSBC (n = 1,385, 48.0%) | LSBC (n = 1,499, 52.0%) | p-value | HSBC (n = 10,849 57.0%) | LSBC (n = 8,170, 43.0%) | p-value |
| ***Comorbidities (n, %)*** | | | | | | | | | |
| *Upper airway resistance syndrome* | 62 (0.6%) | 28 (0.5%) | 1.00 | 6 (0.4%) | 6 (0.4%) | 1.00 | 63 (0.6%) | 39 (0.5%) | 1.00 |
| *Q fever* | 218 (2.2%) | 52 (0.8%) | **<0.001** | 15 (1.1%) | 10 (0.7%) | 1.00 | 229 (2.1%) | 66 (0.8%) | **<0.001** |
| *Bipolar disorder* | 238 (2.4%) | 105 (1.7%) | **0.01** | 33 (2.4%) | 18 (1.2%) | 0.68 | 263 (2.4%) | 131 (1.6%) | **<0.001** |
| *Cancer* | 426 (4.3%) | 388 (6.3%) | **<0.001** | 55 (4.0%) | 67 (4.5%) | 1.00 | 461 (4.2%) | 475 (5.8%) | **<0.001** |
| *Coeliac disease* | 368 (3.7%) | 148 (2.4%) | **<0.001** | 34 (2.5%) | 34 (2.3%) | 1.00 | 393 (3.6%) | 191 (2.3%) | **<0.001** |
| *Diabetes* | 649 (6.5%) | 357 (5.8%) | 1.00 | 123 (8.9%) | 95 (6.3%) | 0.34 | 742 (6.8%) | 482 (5.9%) | 0.33 |
| *Haemochromatosis* | 90 (0.9%) | 51 (0.8%) | 1.00 | 24 (1.7%) | 22 (1.5%) | 1.00 | 110 (1.0%) | 77 (0.9%) | 1.00 |
| ***SYMPTOMS IN PAST 6 MONTHS (n, %)*** | | | | | | | | | |
| *COLD OR FLU-LIKE SYMPTOMS* | | | | | | | | | |
| *Sore throat* | 7608 (76.6%) | 2958 (47.7%) | **<0.001** | 883 (63.8%) | 470 (31.4%) | **<0.001** | 8220 (75.8%) | 3699 (45.3%) | **<0.001** |
| *Flu-like feeling* | 8607 (86.7%) | 3672 (59.2%) | **<0.001** | 1185 (85.6%) | 827 (55.2%) | **<0.001** | 9423 (86.9%) | 4868 (59.6%) | **<0.001** |
| *Fever or chills* | 6063 (60.8%) | 1316 (21.2%) | **<0.001** | 768 (55.5%) | 253 (16.9%) | **<0.001** | 6627 (61.1%) | 1746 (21.4%) | **<0.001** |
| *Swollen or tender neck/armpit glands* | 7051 (71.0%) | 2303 (37.1%) | **<0.001** | 825 (59.6%) | 368 (24.5%) | **<0.001** | 7642 (70.4%) | 2905 (35.6%) | **<0.001** |
| *Viral infections with long recovery* | 4794 (48.3%) | 1748 (28.2%) | **<0.001** | 587 (42.4%) | 359 (23.9%) | **<0.001** | 5214 (48.1%) | 2274 (27.8%) | **<0.001** |
| *Fewer viral infections than before* | 1104 (11.1%) | 661 (10.7%) | 0.38 | 163 (11.8%) | 180 (12.0%) | 0.89 | 1220 (11.2%) | 888 (10.9%) | 0.43 |
| *SENSITIVITIES* | | | | | | | | | |
| *Alcohol* | 5483 (55.2%) | 2270 (36.6%) | **<0.001** | 706 (51.0%) | 479 (32.0%) | **<0.001** | 5969 (55.0%) | 2969 (36.3%) | **<0.001** |
| *Chemicals* | 4171 (42.0%) | 1152 (18.6%) | **<0.001** | 437 (31.6%) | 169 (11.3%) | **<0.001** | 4497 (41.5%) | 1432 (17.5%) | **<0.001** |
| *Food* | 6415 (64.6%) | 2168 (35.0%) | **<0.001** | 765 (55.2%) | 348 (23.2%) | **<0.001** | 6983 (64.4%) | 2713 (33.2%) | **<0.001** |
| *Medicine* | 3586 (36.1%) | 852 (13.7%) | **<0.001** | 372 (26.9%) | 151 (10.1%) | **<0.001** | 3882 (35.8%) | 1079 (13.2%) | **<0.001** |
| *Smells* | 6858 (69.0%) | 1872 (30.2%) | **<0.001** | 614 (44.3%) | 247 (16.5%) | **<0.001** | 7305 (67.3%) | 2286 (28.0%) | **<0.001** |
| *Other things* | 2485 (25.0%) | 700 (11.3%) | **<0.001** | 367 (26.5%) | 212 (14.1%) | **<0.001** | 2786 (25.7%) | 978 (12.0%) | **<0.001** |
| *Light* | 8185 (82.4%) | 3185 (51.3%) | **<0.001** | 1075 (77.6%) | 611 (40.8%) | **<0.001** | 9080 (83.7%) | 3976 (48.7%) | **<0.001** |
| *Noise* | 9107 (91.7%) | 4458 (71.9%) | **<0.001** | 1195 (86.3%) | 889 (59.3%) | **<0.001** | 9944 (91.7%) | 5705 (69.8%) | **<0.001** |
| *Touch* | 6197(62.4%) | 1293 (20.8%) | **<0.001** | 585 (42.2%) | 199 (13.3%) | **<0.001** | 6674 (61.5%) | 1600 (19.6%) | **<0.001** |
| *HEADACHES* | | | | | | | | | |
| *Headache* | 8828 (88.9%) | 3853 (62.1%) | **<0.001** | 1192 (86.1%) | 811 (54.1%) | **<0.001** | 9672 (89.2%) | 5012 (61.3%) | **<0.001** |
| *Migraine* | 5590 (56.3%) | 1432 (23.1%) | **<0.001** | 605 (43.7%) | 234 (15.6%) | **<0.001** | 6065 (55.9%) | 1796 (22.0%) | **<0.001** |
| *Eye pain or pain behind eyes* | 7762 (78.2%) | 2182 (35.2%) | **<0.001** | 1089 (78.6%) | 467 (31.2%) | **<0.001** | 8600 (79.3 %) | 2900 (35.5%) | **<0.001** |
| *Feeling of head/base of skull pressure* | 8395 (84.5%) | 2878 (46.4%) | **<0.001** | 1137 (82.1%) | 609 (40.6%) | **<0.001** | 9211 (84.9%) | 3808 (46.6%) | **<0.001** |

HSBC = high symptom burden cluster, LSBC = lower symptom burden cluster. P-values < 0.05 are shown in bold.

| Characteristic | Females (n = 16,135) | | | Males (n = 2,884) | | | Total (n = 19,019) | | |
|---|---|---|---|---|---|---|---|---|---|
| | **HSBC (n = 9,932, 61.6%)** | **LSBC (n = 6,203, 38.4%)** | **p-value** | **HSBC (n = 1,385, 48.0%)** | **LSBC (n = 1,499, 52.0%)** | **p-value** | **HSBC (n = 10,849 57.0%)** | **LSBC (n = 8,170, 43.0%)** | **p-value** |
| ***SYMPTOMS IN PAST 6 MONTHS (n, %)*** | | | | | | | | | |
| *GUT SYMPTOMS* | | | | | | | | | |
| *Feeling sick (nausea)* | 7921 (79.8%) | 2261 (36.5%) | **<0.001** | 945 (68.2%) | 330 (22.0%) | **<0.001** | 8600 (79.3%) | 2857 (35.0%) | **<0.001** |
| *Gut or irritable bowel-type symptoms* | 9427 (94.9%) | 4730 (76.3%) | **<0.001** | 1265 (91.3%) | 970 (64.7%) | **<0.001** | 10287 (94.8%) | 6105 (74.7%) | **<0.001** |
| *Heartburn* | 5813 (58.5%) | 1716 (28.4%) | **<0.001** | 855 (61.7%) | 380 (25.4%) | **<0.001** | 6476 (59.7%) | 2333 (28.6%) | **<0.001** |
| *MUSCLE/JOINT SYMPTOMS* | | | | | | | | | |
| *Muscle pain* | 9505 (95.7%) | 4761 (76.8%) | **<0.001** | 1290 (93.1%) | 1038 (69.2%) | **<0.001** | 10397 (95.8%) | 6197 (75.9%) | **<0.001** |
| *Muscle twitching/spasms* | 8079 (81.3%) | 2447 (39.4%) | **<0.001** | 1133 (81.8%) | 623 (41.6%) | **<0.001** | 8903 (82.1%) | 3379 (41.4%) | **<0.001** |
| *Muscle stiffness* | 8733 (87.9%) | 3737 (60.2%) | **<0.001** | 1211 (87.4%) | 818 (54.6%) | **<0.001** | 9586 (88.4%) | 4913 (60.1%) | **<0.001** |
| *Joint pain with no swelling or redness* | 7295 (73.4%) | 3058 (49.3%) | **<0.001** | 1005 (72.6%) | 661 (44.1%) | **<0.001** | 8046 (74.2%) | 3973 (48.6%) | **<0.001** |
| *Migratory joint pain (no swell/no red)* | 7250 (73.0%) | 2167 (34.9%) | **<0.001** | 908 (65.6%) | 388 (25.9%) | **<0.001** | 7899 (72.8%) | 2814 (34.4%) | **<0.001** |
| *Muscle weakness* | 9179 (92.4%) | 4226 (68.1%) | **<0.001** | 1275 (92.1%) | 997 (66.5%) | **<0.001** | 10062 (92.7%) | 5615 (68.7%) | **<0.001** |
| *Chest pain* | 5305 (53.4%) | 1055 (17.0%) | **<0.001** | 786 (56.8%) | 247 (16.5%) | **<0.001** | 5973 (55.1%) | 1420 (17.4%) | **<0.001** |
| *PROBLEMS WITH THINKING, SENSATION, OR MOVEMENT* | | | | | | | | | |
| *Temporary eyesight/focus problems* | 6936 (69.8%) | 1676 (27.0%) | **<0.001** | 1023 (73.9%) | 441 (29.4%) | **<0.001** | 7732 (71.3%) | 2344 (28.7%) | **<0.001** |
| *Numbness or tingling in arms or legs* | 7505 (75.6%) | 2101 (33.9%) | **<0.001** | 1058 (76.4%) | 425 (28.4%) | **<0.001** | 8278 (76.3%) | 2811 (34.4%) | **<0.001** |
| *Poor standing balance/steadiness* | 8876 (89.4%) | 2992 (48.2%) | **<0.001** | 1213 (87.6%) | 635 (42.4%) | **<0.001** | 9738 (89.8%) | 3978 (48.7%) | **<0.001** |
| *Poor walking coordination/steadiness* | 7918 (79.7%) | 2105 (33.9%) | **<0.001** | 1081 (78.1%) | 447 (29.8%) | **<0.001** | 8726 (80.4%) | 2825 (34.6%) | **<0.001** |
| *Short-term memory problems* | 8217 (82.7%) | 3336 (53.8%) | **<0.001** | 1189 (85.8%) | 876 (58.4%) | **<0.001** | 9068 (83.6%) | 4550 (55.7%) | **<0.001** |
| *Disorientation* | 6543 (65.9%) | 1440 (23.2%) | **<0.001** | 967 (69.8%) | 360 (24.0%) | **<0.001** | 7294 (67.2%) | 2016 (24.7%) | **<0.001** |
| *Hard to understand things/think clearly* | 8808 (88.7%) | 3995 (64.4%) | **<0.001** | 1230 (88.8%) | 943 (62.9%) | **<0.001** | 9684 (89.3%) | 5292 (64.8%) | **<0.001** |
| *Problems finding or saying words* | 9343 (94.1%) | 4805 (77.5%) | **<0.001** | 1260 (91.0%) | 1022 (68.2%) | **<0.001** | 10200 (94.0%) | 6230 (76.3%) | **<0.001** |
| *Hard to remember things* | 9387 (94.5%) | 4857 (78.3%) | **<0.001** | 1307 (94.4%) | 1138 (75.9%) | **<0.001** | 10283 (94.8%) | 6406 (78.4%) | **<0.001** |
| *Slow thinking* | 8782 (88.4%) | 4069 (65.6%) | **<0.001** | 1248 (90.1%) | 1014 (67.6%) | **<0.001** | 9669 (89.1%) | 5444 (66.6%) | **<0.001** |
| *Hard to make decisions* | 8796 (88.6%) | 4154 (67.0%) | **<0.001** | 1223 (88.3%) | 974 (65.0%) | **<0.001** | 9660 (89.0%) | 5487 (67.2 %) | **<0.001** |
| *Speech problems* | 5962 (60.0%) | 1188 (19.2%) | **<0.001** | 816 (58.9%) | 268 (17.9%) | **<0.001** | 6604 (60.9%) | 1630 (20.0%) | **<0.001** |
| *Ringing in ears (Tinnitus)* | 6452 (65.0%) | 2209 (35.6%) | **<0.001** | 937 (67.7%) | 597 (39.8%) | **<0.001** | 7160 (66.0%) | 3035 (37.1%) | **<0.001** |
| *SLEEP* | | | | | | | | | |
| *Problems with sleep quality or length* | 8961 (90.2%) | 4667 (75.2%) | **<0.001** | 1258 (90.8%) | 1092 (72.8%) | **<0.001** | 9825 (90.6%) | 6153 (75.3%) | **<0.001** |
| *Night sweats* | 6716 (67.6%) | 2250 (36.3%) | **<0.001** | 860 (62.1%) | 403 (26.9%) | **<0.001** | 7338 (67.6%) | 2891 (35.4%) | **<0.001** |
| *Feeling more sleepy than is normal* | 8686 (87.5%) | 4163 (67.1%) | **<0.001** | 1194 (86.2%) | 1021 (68.1%) | **<0.001** | 9517 (87.7%) | 5547 (67.9%) | **<0.001** |

HSBC = high symptom burden cluster, LSBC = lower symptom burden cluster. P-values < 0.05 are shown in bold.

| Characteristic | Females (n = 16,135) | | | Males (n = 2,884) | | | Total (n = 19,019) | | |
|---|---|---|---|---|---|---|---|---|---|
| | **HSBC (n = 9,932, 61.6%)** | **LSBC (n = 6,203, 38.4%)** | **p-value** | **HSBC (n = 1,385, 48.0%)** | **LSBC (n = 1,499, 52.0%)** | **p-value** | **HSBC (n = 10,849 57.0%)** | **LSBC (n = 8,170, 43.0%)** | **p-value** |
| ***SYMPTOMS IN PAST 6 MONTHS (n, %)*** | | | | | | | | | |
| *NEUROENDOCRINE SYMPTOMS* | | | | | | | | | |
| *Symptoms worse with stress* | 9086 (91.5%) | 4821 (77.7%) | **<0.001** | 1230 (88.8%) | 1071 (71.4%) | **<0.001** | 9913 (91.4%) | 6295 (77.1%) | **<0.001** |
| *Difficulty coping with being in very hot or cold places* | 8955 (90.2%) | 4028 (64.9%) | **<0.001** | 1149 (83.0%) | 756 (50.4%) | **<0.001** | 9759 (90.0%) | 5129 (62.8%) | **<0.001** |
| *Less interest in sex and/or sexual function difficulties* | 7084 (71.3%) | 2934 (47.3%) | **<0.001** | 998 (72.1%) | 718 (47.9%) | **<0.001** | 7789 (71.8%) | 3945 (48.3%) | **<0.001** |
| *Unusual change in appetite or weight* | 7725 (77.8%) | 2548 (41.1%) | **<0.001** | 990 (71.5%) | 456 (30.4%) | **<0.001** | 8426 (77.7%) | 3293 (40.3%) | **<0.001** |
| *AUTOMATIC BODY FUNCTIONS* | | | | | | | | | |
| *Difficulty remaining standing* | 8318 (83.7%) | 3332 (53.7%) | **<0.001** | 1089 (78.6%) | 580 (38.7%) | **<0.001** | 9089 (83.8%) | 4230 (51.8%) | **<0.001** |
| *Dizzy or faint when standing* | 8445 (85.0%) | 2635 (42.5%) | **<0.001** | 1177 (85.0%) | 562 (37.5%) | **<0.001** | 9308 (85.8%) | 3511 (43.0%) | **<0.001** |
| *Palpitations while standing/other times* | 7307 (73.6%) | 2050 (33.0%) | **<0.001** | 904 (65.3%) | 322 (21.5%) | **<0.001** | 7996 (73.7%) | 2587 (31.7%) | **<0.001** |
| *Light-headedness* | 8755 (88.1%) | 2776 (44.8%) | **<0.001** | 1199 (86.6%) | 569 (38.0%) | **<0.001** | 9605 (88.5%) | 3694 (45.2%) | **<0.001** |
| *Having a very pale face* | 5437 (54.7%) | 1479 (23.8%) | **<0.001** | 558 (40.3%) | 254 (16.9%) | **<0.001** | 5877 (54.2%) | 1851 (22.7%) | **<0.001** |
| *Excessive sweating* | 5857 (59.0%) | 1420 (22.9%) | **<0.001** | 762 (55.0%) | 307 (20.5%) | **<0.001** | 6430 (59.3%) | 1916 (23.5%) | **<0.001** |
| *Cold hands or feet* | 7484 (75.4%) | 3255 (52.5%) | **<0.001** | 976 (70.5%) | 522 (34.8%) | **<0.001** | 8293 (76.4%) | 3944 (48.3%) | **<0.001** |
| *Bladder problems* | 7038 (70.9%) | 2444 (39.4%) | **<0.001** | 921 (66.5%) | 574 (38.3%) | **<0.001** | 7699 (71.0%) | 3278 (40.1%) | **<0.001** |
| *Breathing difficulty during activity/effort* | 7041 (70.9%) | 1819 (29.3%) | **<0.001** | 963 (69.5%) | 375 (25.0%) | **<0.001** | 7774 (71.7%) | 2424 (29.7%) | **<0.001** |
| *Feeling of being sick or unwell* | 8355 (84.1%) | 2579 (41.6%) | **<0.001** | 1067 (77.0%) | 479 (32.0%) | **<0.001** | 9128 (84.1%) | 3352 (41.0%) | **<0.001** |
| *Tight feeling in the chest* | 4878 (49.1%) | 986 (15.9%) | **<0.001** | 738 (53.3%) | 192 (12.8%) | **<0.001** | 5594 (51.6%) | 1200 (14.7%) | **<0.001** |
| *Feeling of burning in the lungs* | 1340 (13.5%) | 147 (2.4%) | **<0.001** | 229 (16.5%) | 39 (2.6%) | **<0.001** | 1558 (14.4%) | 197 (2.4%) | **<0.001** |
| *MOOD* | | | | | | | | | |
| *Feeling easily anxious or nervous* | 8232 (82.9%) | 3705 (59.7%) | **<0.001** | 1153 (83.2%) | 827 (55.2%) | **<0.001** | 9024 (83.2%) | 4894 (59.9%) | **<0.001** |
| *Feeling low or down* | 8677 (87.4%) | 4523 (72.9%) | **<0.001** | 1242 (89.7%) | 1093 (72.9%) | **<0.001** | 9537 (87.9%) | 5998 (73.4%) | **<0.001** |
| *Feeling easily annoyed or irritable* | 8136 (81.9%) | 3837 (61.9%) | **<0.001** | 1209 (87.3%) | 1070 (71.4%) | **<0.001** | 8993 (82.9%) | 5259 (64.4%) | **<0.001** |
| *Mood swings* | 6177 (62.2%) | 1720 (27.7%) | **<0.001** | 922 (66.6%) | 454 (30.3%) | **<0.001** | 6863 (63.3%) | 2410 (29.5%) | **<0.001** |
| *Racing thoughts* | 6441 (64.9%) | 1989 (32.1%) | **<0.001** | 948 (68.4%) | 476 (31.8%) | **<0.001** | 7144 (65.8%) | 2710 (33.2%) | **<0.001** |
| *Feeling worried* | 7529 (75.8%) | 2999 (48.3%) | **<0.001** | 1059 (76.5%) | 653 (43.6%) | **<0.001** | 8306 (76.6%) | 3934 (48.2%) | **<0.001** |

HSBC = high symptom burden cluster, LSBC = lower symptom burden cluster. P-values < 0.05 are shown in bold

**Supplemental Table 3.** Three associations with sub-genome wide significance ($p < 8 \times 10^{-7}$) of association to HSBC status.

| **Chromosome: position (GRCh38) alleles** | **Variant tested, rsID** | **Nearest Protein Coding Gene(s)** | **p-value, effect size (b), allele frequency (AF)** | **Gene function annotation** |
|---|---|---|---|---|
| **15: 60,884,050 C/G** | rs341388 | *RORA* | $2.0 \times 10^{-7}$ b = +0.219 AF = 0.341 | Chronobiology, inflammation, type 2 innate lymphoid cell development |
| **7: 150,346,134 T/A** | rs883139 | *RARRES2* | $5.8 \times 10^{-7}$ b =-0.333 AF = 0.097 | Adipokine, immune response, antimicrobial, anti-inflammation. |
| **2: 234,081,189 A/G** | rs60995367 | *SPP2* / *TRPM8* | $7.59 \times 10^{-7}$ b = -0.499 AF = 0.038 | *SPP2* (bone metabolism; liver); *TRPM8* (temperature regulation, pain sensing, migraine) |

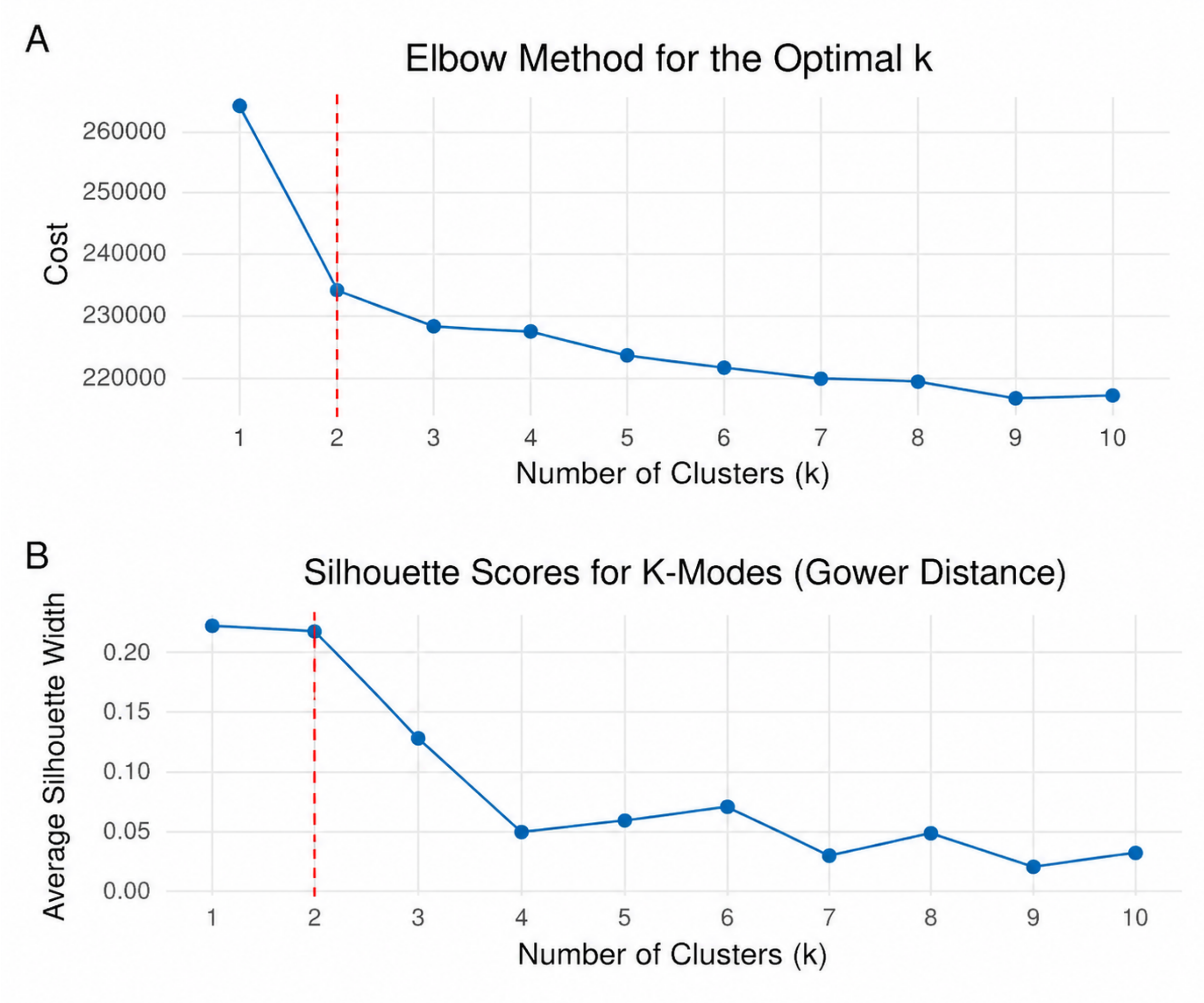

**Supplemental Figure 1.** Cluster metrics for determining the optimal number of clusters for the total sample. A) Elbow method for determining the optimal number of clusters, k. The elbow in the plot is the point at which increasing k results in diminishing returns in cost reduction (reducing the within-cluster dissimilarity between points). The vertical dashed red line indicates the optimal number of clusters. B) Average silhouette scores for k clusters to determine the optimal k. Higher average silhouette scores indicate better cluster quality. The vertical dashed red line indicates the optimal number of clusters.

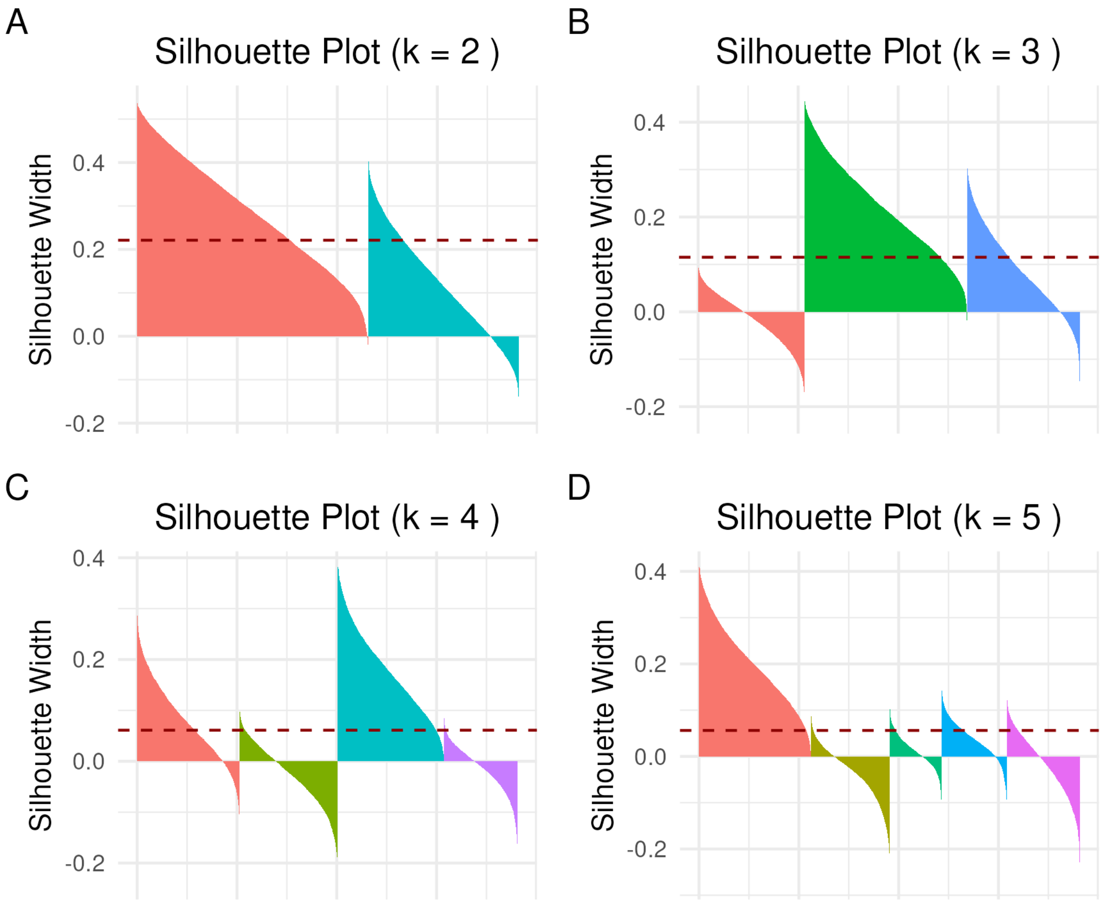

**Supplemental Figure 2.** Silhouette plots for the total sample. The plots show the silhouette width when k = 2 (A), k = 3 (B), k = 4 (C), and k = 5 (D). Higher silhouette width indicates more positive silhouette values and a higher degree of cohesion between data points within a cluster, and better separation between the clusters. Negative silhouette values indicate those points are more similar to another cluster than their own cluster based on the distance measure used (Hamming distance). The average silhouette width is indicated by the dashed horizontal line.

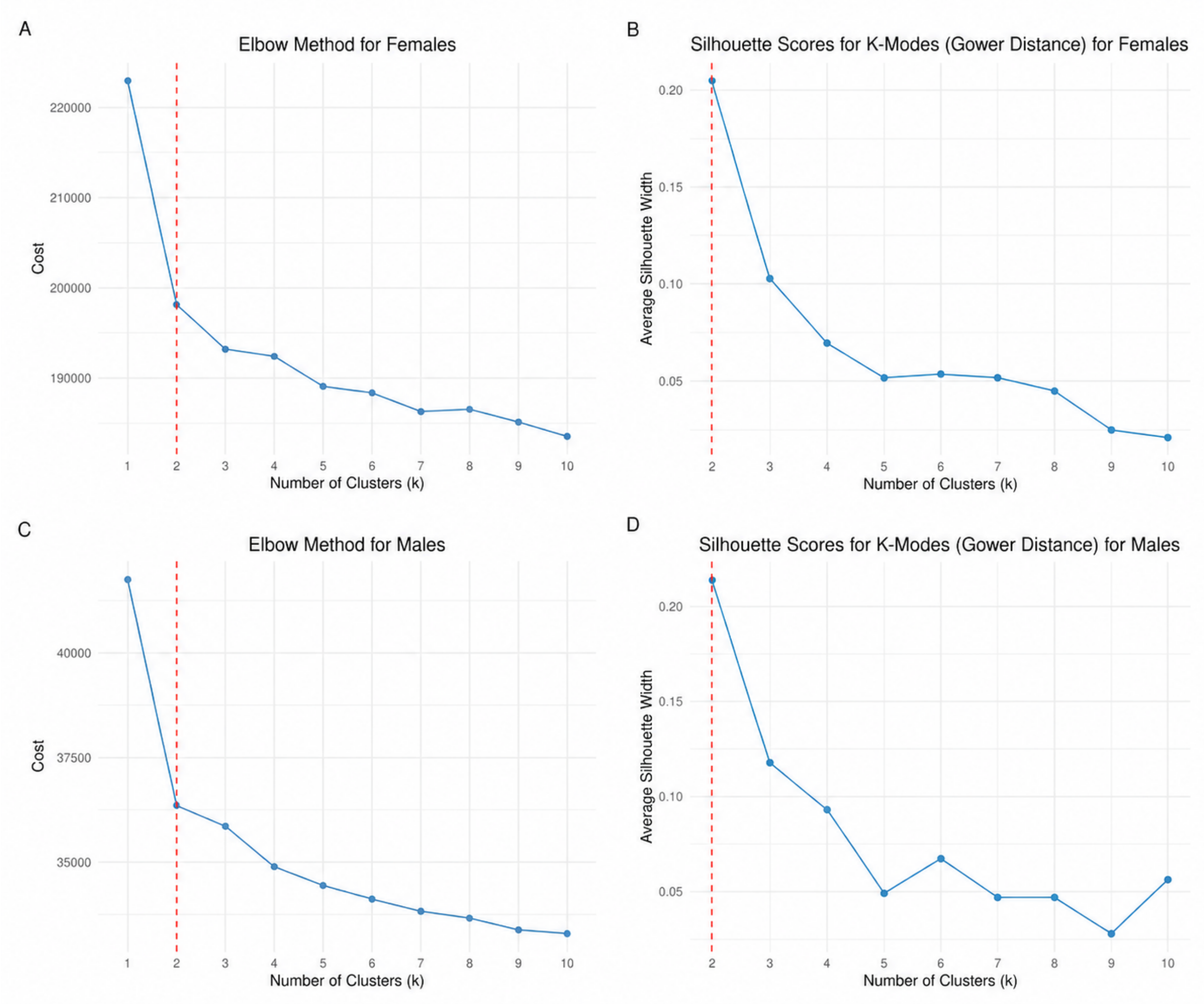

**Supplemental Figure 3.** Cluster metrics for determining the optimal number of clusters for sex-based subgroups. A, C) Elbow method for determining the optimal number of clusters for females (A) and men (C). The elbow in the plot is the point at which increasing k results in diminishing returns in cost reduction (reducing the within-cluster dissimilarity between points). The vertical dashed red line indicates the optimal number of clusters. B, D) Average silhouette scores for k clusters to determine the optimal k for females (B) and men (D). Higher average silhouette scores indicate better cluster quality. The vertical dashed red line indicates the optimal number of clusters.

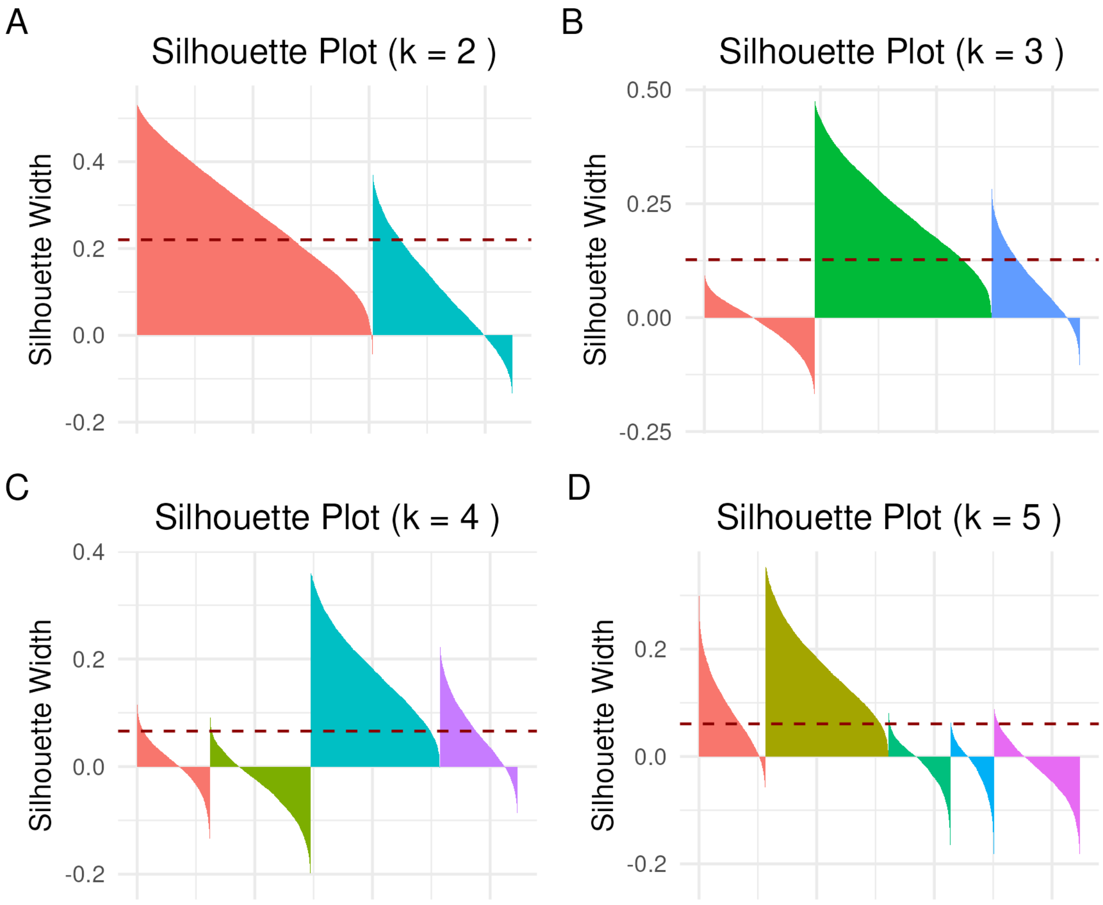

**Supplemental Figure 4.** Silhouette plots for females. The plots show the silhouette width when k = 2 (A), k = 3 (B), k = 4 (C), and k = 5 (D). Higher silhouette width indicates more positive silhouette values and a higher degree of cohesion between data points within a cluster, and better separation between the clusters. Negative silhouette values indicate those points are more similar to another cluster than their own cluster based on the distance measure used (Hamming distance). The average silhouette width is indicated by the dashed horizontal line.

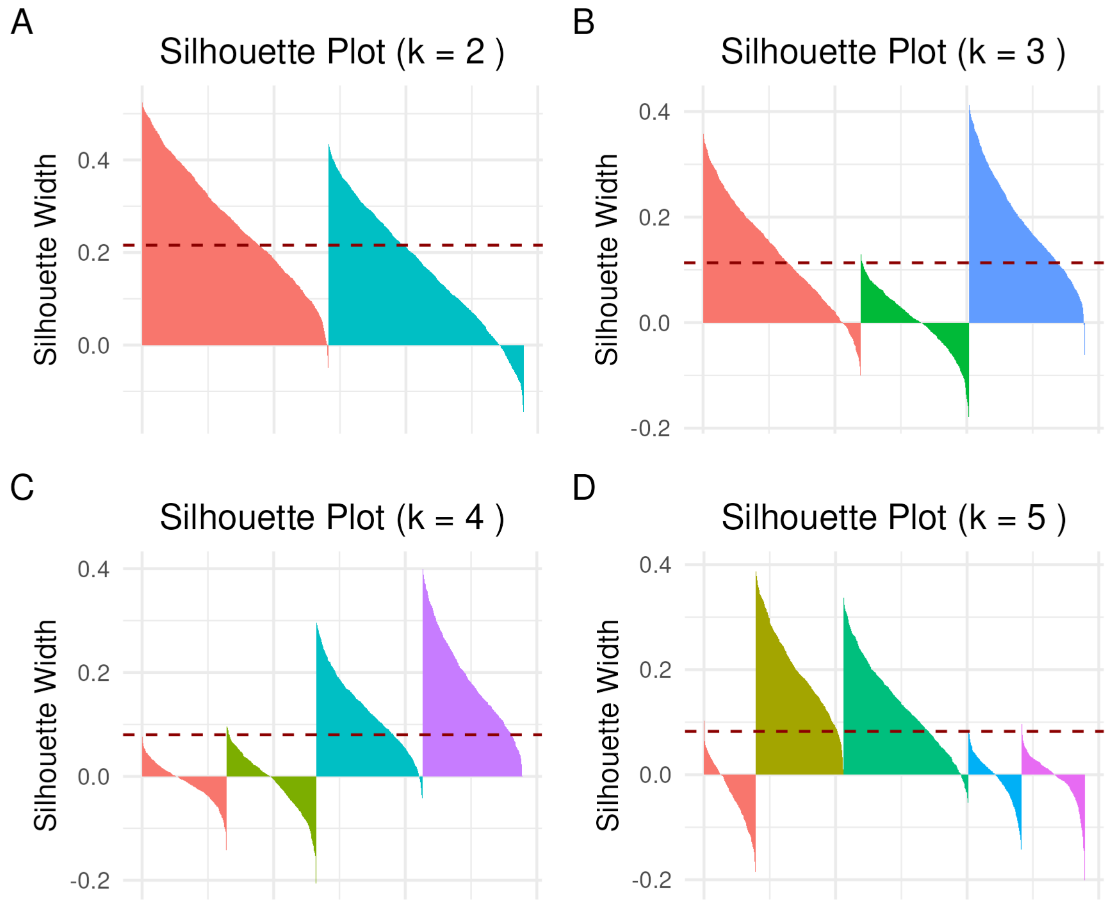

**Supplemental Figure 5.** Silhouette plots for males. The plots show the silhouette width when k = 2 (A), k = 3 (B), k = 4 (C), and k = 5 (D). Higher silhouette width indicates more positive silhouette values and a higher degree of cohesion between data points within a cluster, and better separation between the clusters. Negative silhouette values indicate those points are more similar to another cluster than their own cluster based on the distance measure used (Hamming distance). The average silhouette width is indicated by the dashed horizontal line.

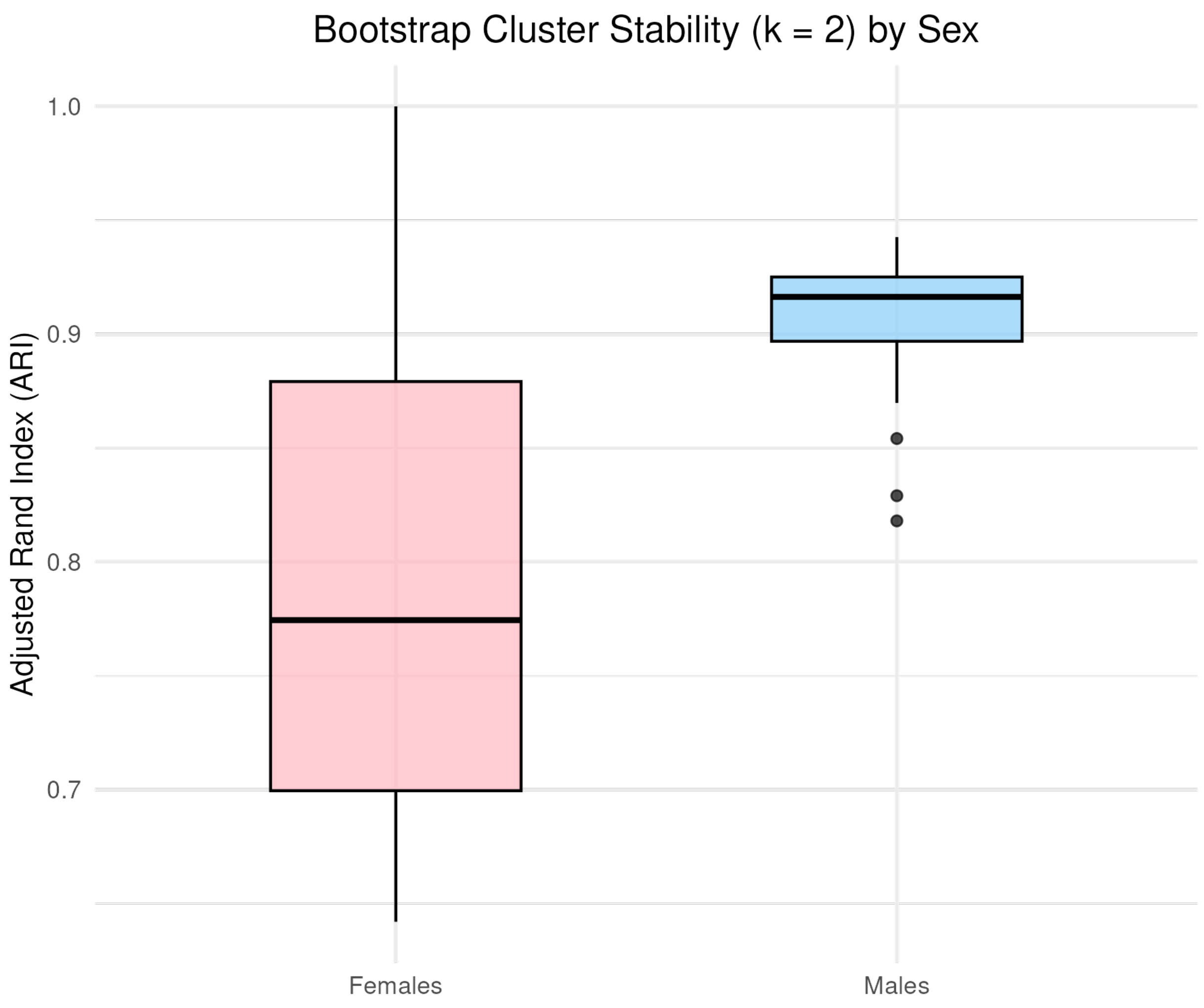

**Supplemental Figure 6.** Box-and-whisker plot showing the distribution of Adjusted Rand Index values for k = 2 clusters, stratified by sex. ARI values range from 0 - 1, with higher values indicating better cluster stability. ARI values are calculated from 100 bootstrap replicates.
